# ECG Criteria for Left Ventricular Hypertrophy in Hypertensive Black Africans: Insights from the CoArtHA Trial

**DOI:** 10.1101/2025.07.31.25332550

**Authors:** Annina Vischer, Valeriya Nemtsova, Blaise Lukau, Martin Rohacek, Jonathan Macko, Johanna Oehri, Herry Mapesi, Fiona Vanobberghen, Andrew Katende, Ravi Gupta, Herieth Ismael Wilson, Alain Amstutz, Josephine Muhairwe, Elizabeth Senkoro, Theonestina Byakuzana, Geofrey Mbunda, Jamali Siru, Winfrid Gingo, Niklaus Daniel Labhardt, Maja Weisser, Thilo Burkard

## Abstract

**Background:** Electrocardiographic (ECG) criteria for detecting left ventricular hypertrophy (LVH) were derived largely from non-African populations. Whether these indices accurately identify LVH in African adults with untreated hypertension remains uncertain.

**Objective:** To evaluate the diagnostic accuracy of established ECG-LVH criteria compared with echocardiographic left ventricular mass index (LVMI) in Black African adults with untreated, uncomplicated hypertension enrolled in the CoArtHA trial.

**Methods:** In this subanalysis, we included participants from rural Tanzania and Lesotho who underwent baseline 12-lead ECG and focused transthoracic echocardiography (fTTE). LVH was defined as LVMI > 95 g/m² in women and > 115 g/m² in men. Diagnostic performance of multiple ECG-LVH criteria was assessed using Spearman correlation, sensitivity, specificity, and area under the receiver-operating characteristic curve (AUC).

**Results:** Echocardiographic LVH was present in 56 participants (5%). Among continuous ECG indices, the Cornell voltage product adjusted by + 0.8 mV in women showed the highest correlation with LVMI (rho = 0.373, p < 0.001) and achieved the best overall discrimination (AUC = 0.719). Standard cut-offs provided a specificity of 79.5% and sensitivity of 64.3%, whereas an adapted cut-off of 277.8 mV*ms increased specificity above 90%. Other indices, including MESA- and Sokolow-Lyon-based criteria, showed limited accuracy.

**Conclusions:** In this large cohort of hypertensive Black African adults, the Cornell voltage product (with + 0.8 mV adjustment for women) demonstrated the best diagnostic performance for detecting LVH compared with echocardiography. These results highlight the need to verify the diagnostic validity of established ECG criteria in African populations and support their pragmatic use to guide cardiovascular risk stratification in low-resource settings.

**Trial registration:** Clinicaltrials.gov NCT04129840. Registered on 17 October 2019 (https://www.clinicaltrials.gov/).

## INTRODUCTION

Left ventricular hypertrophy (LVH) is a recognized complication of hypertension and associated with a worse prognosis, independent of ethnicity(1). While cardiac magnetic resonance (CMR) is considered the reference method for assessing LV mass, its availability in low-resource settings is limited. Transthoracic echocardiography (TTE) remains the preferred modality in clinical practice, yet even this is often inaccessible in rural regions of sub-Saharan Africa (2, 3). In contrast, electrocardiography (ECG) is inexpensive and widely available, but its diagnostic accuracy for LVH varies considerably (4–6). Most ECG-LVH criteria were developed in predominantly Caucasian populations, with limited validation among African adults (7, 8). Only few studies have evaluated the suitability of the ECG criteria for LVH assessment in adult black African populations (5, 9). In the MESA (the Multi-Ethnic Study of Atherosclerosis) study, the MESA ECG-LVH index was the best at diagnosing LVH in the subgroup of African Americans when compared to CMR (10).

The CoArtHA trial (Identifying the most effective Treatment Strategies to Control Arterial Hypertension in sub-Saharan Africa) enrolled treatment-naïve adults with uncomplicated hypertension in Tanzania and Lesotho (11, 12). Using this unique dataset, we assessed the diagnostic performance of numerous established ECG-LVH criteria against focused echocardiographic (fTTE) LV mass index (LVMI) and explored optimal cut-off values for this population.

## METHODS

### Study design and population

The CoArtHA trial was an investigator-initiated, open-label, randomized controlled trial conducted in rural Tanzania and Lesotho to compare three antihypertensive treatment strategies. Detailed methods and eligibility criteria have been published previously (11, 12). In brief, Black African adults (≥18 years) with untreated, uncomplicated hypertension were enrolled. Key exclusion criteria were symptomatic hypertension, recent cardiovascular events, severe comorbidities, pregnancy, or inability to attend follow-up. This subanalysis included all participants with interpretable baseline ECG and fTTE.

All participants gave written informed consent for the collection, storage and use of clinical data and sample specimens for research purposes within the CoArtHA trial. The study protocol of the CoArtHA trial was approved by the Institutional review board of the Ifakara Health Institute, the National Health Research Ethics Committee in Tanzania, the Tanzania Medicines and Medical Devices Authority, the National Health Research and Ethics Committee and Ministry of Health of Lesotho, as well as the Ethics committee of Northwest and Central Switzerland (11, 12).

This was an exploratory analysis of the CoArtHA trial using fTTE and ECG data from baseline investigations. Patients for whom ECG or fTTE data were not available or of insufficient quality, were excluded (Figure 2).

**Figure 1:**
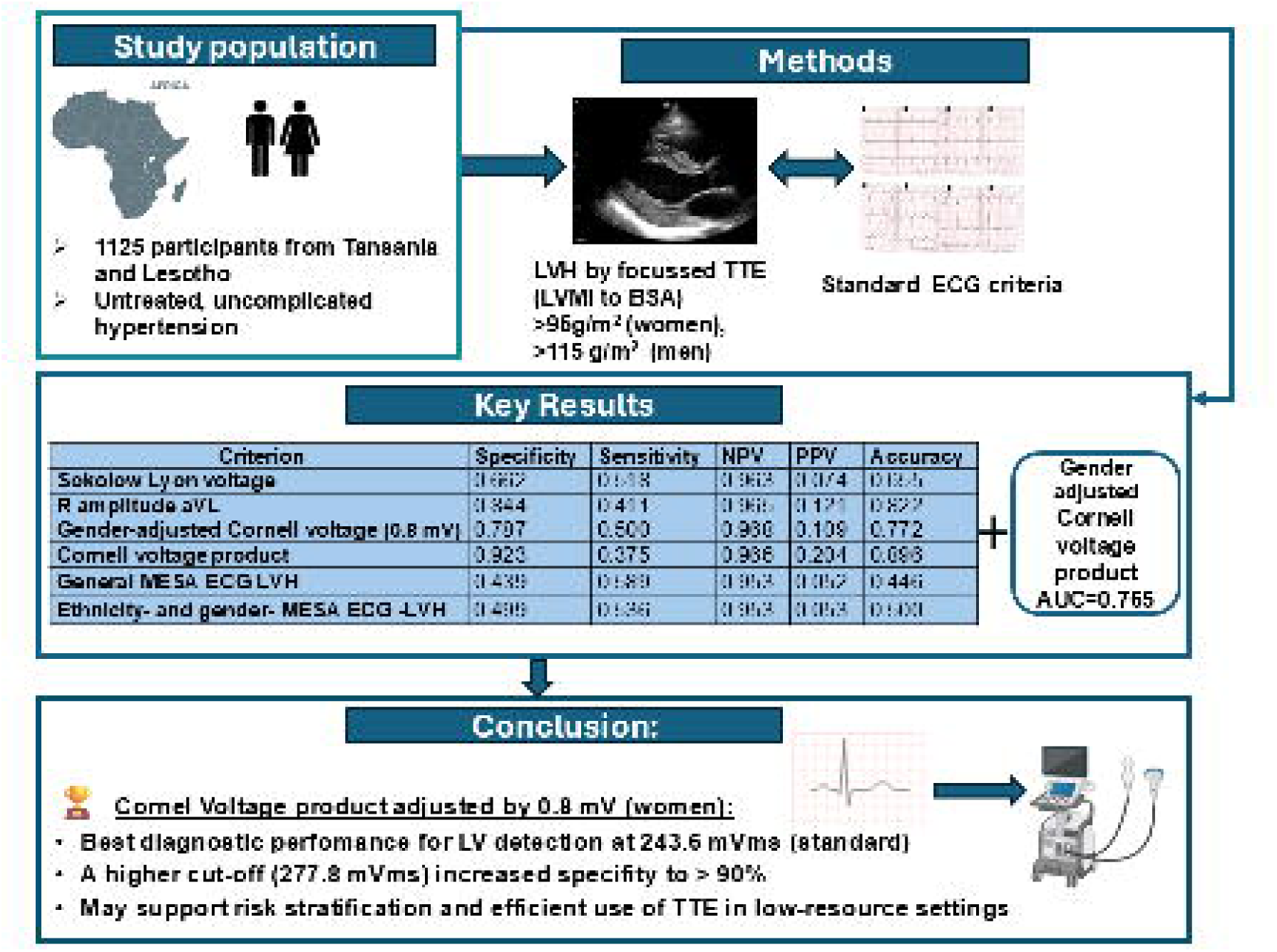
Graphical abstract.

**Figure 2:**
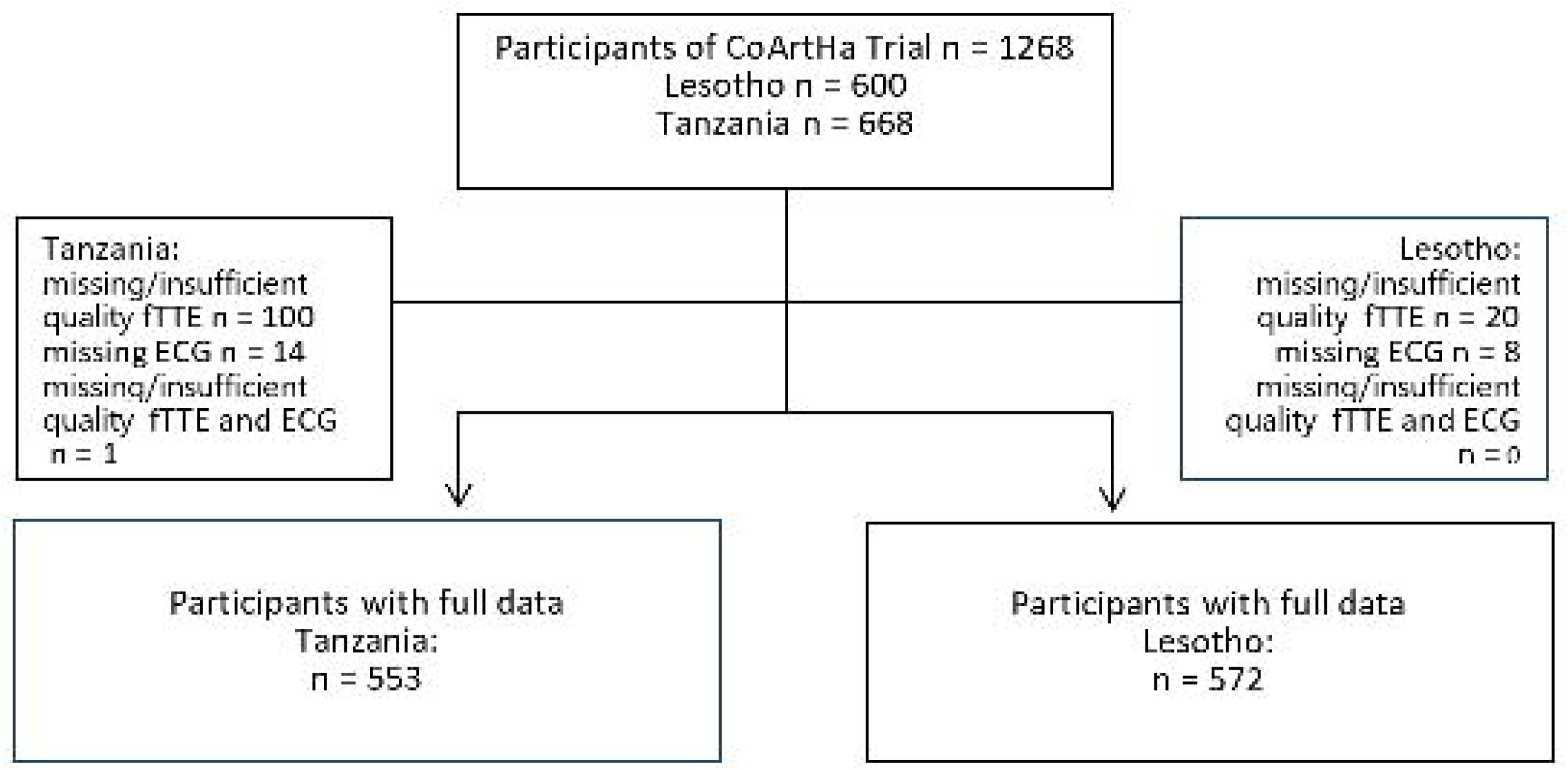
Study flow chart and selection of participants. Legend: fTTE - focused echocardiography, ECG-electrocardiography.

### Study procedures

#### Blood pressure measurement

Blood pressure was measured according to ESC/ESH 2018 guidelines using an automated device (Omron M6 Comfort, HEM-7321-E). Hypertension was defined as systolic ≥ 140 mmHg and/or diastolic ≥ 90 mmHg. The average of the last two of three measurements was used for analysis.(13, 14).

#### Electrocardiography

A 12-lead ECG was recorded at baseline using a Schiller AT-1 electrocardiogram machine at 10 mm/mV calibration and at a speed of 25 mm/s.. All ECGs were printed out including an automated matrix analysis of times and amplitudes and were verified by trained investigators under cardiologist supervision. We compared several established ECG-LVH indices with definitions and thresholds summarized in Table 1). Parameters were analyzed as both dichotomous and continuous variables to evaluate correlation and discriminative ability versus echocardiographic LVH as appropriate.

**Table 1:**
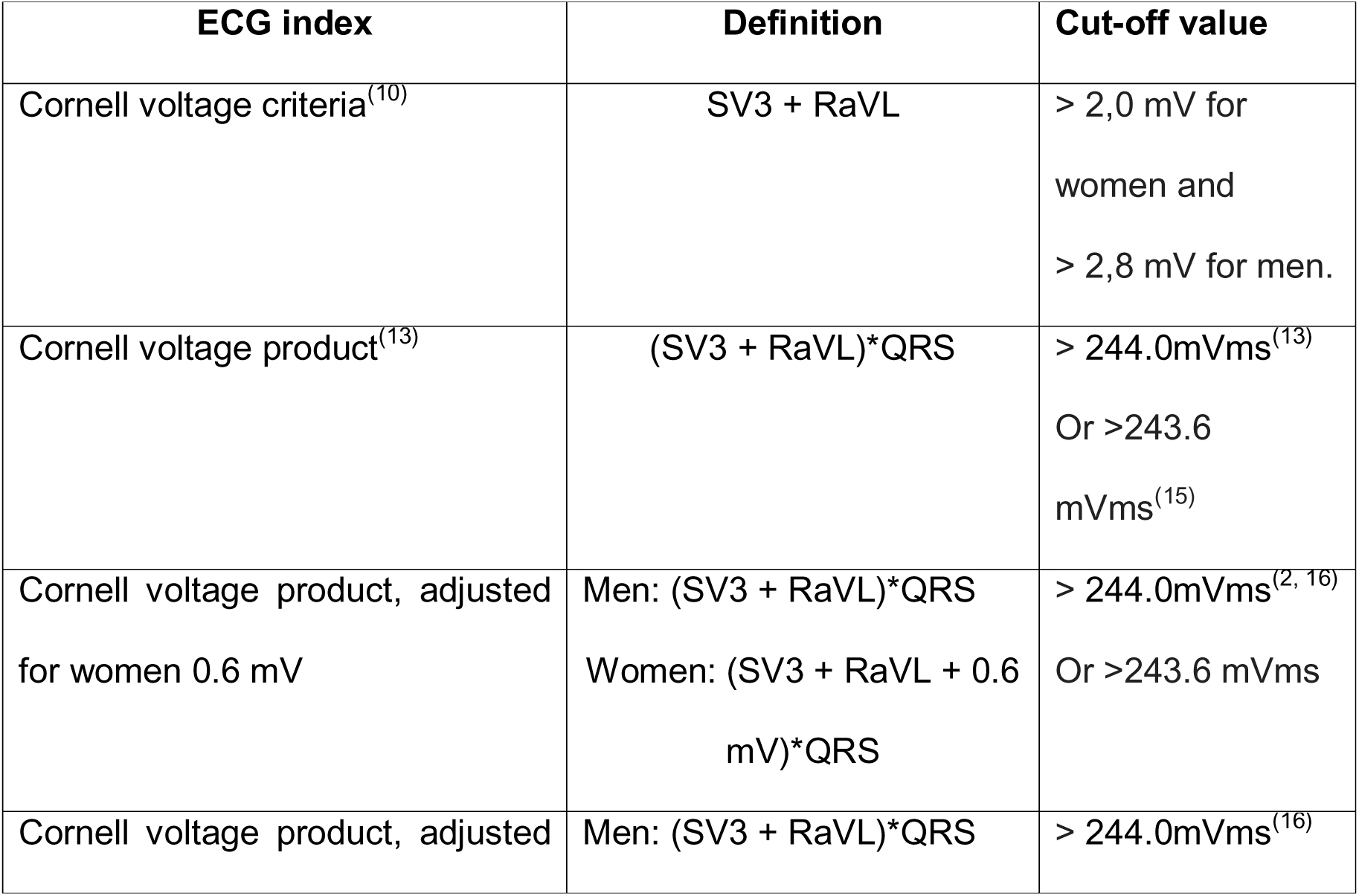
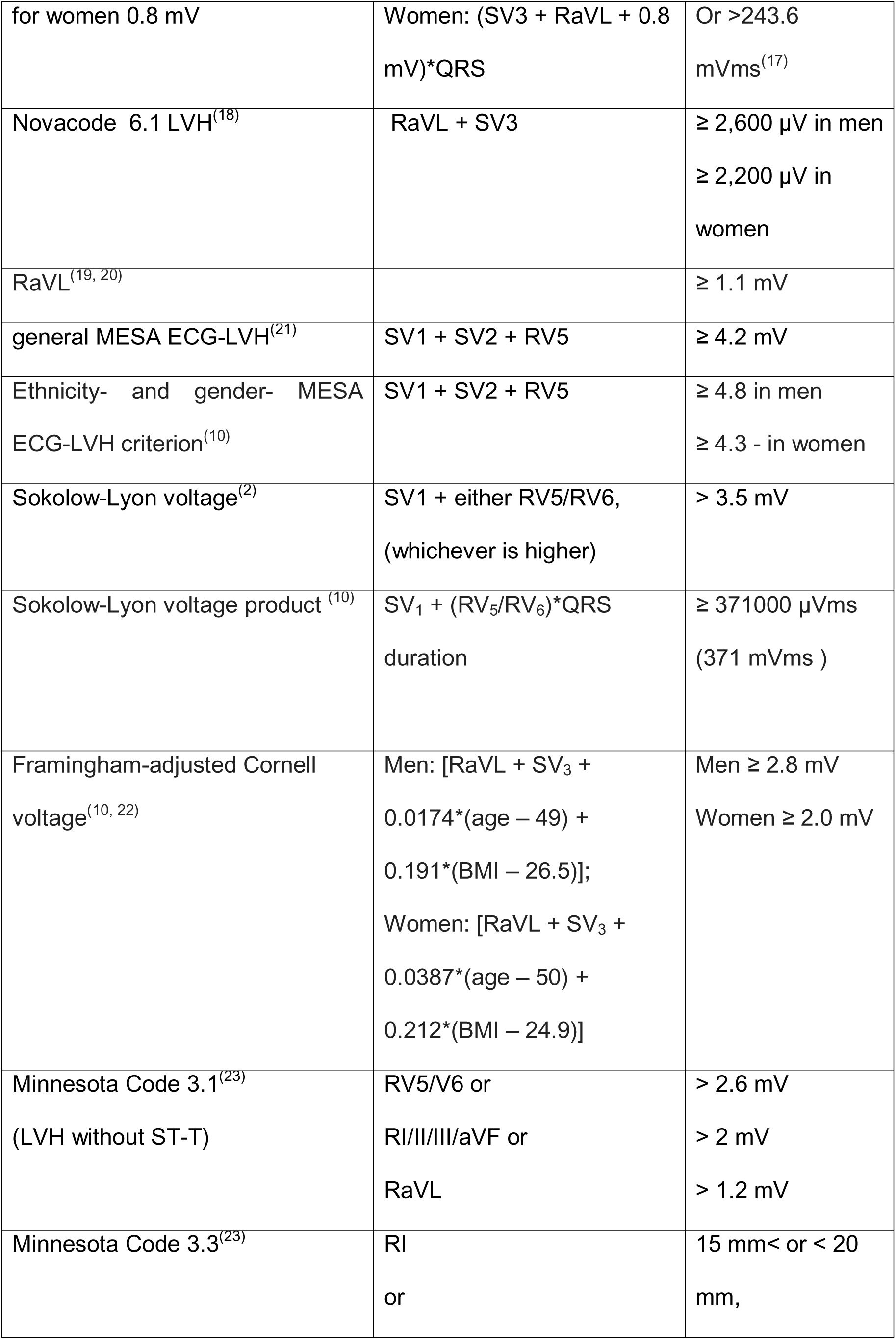
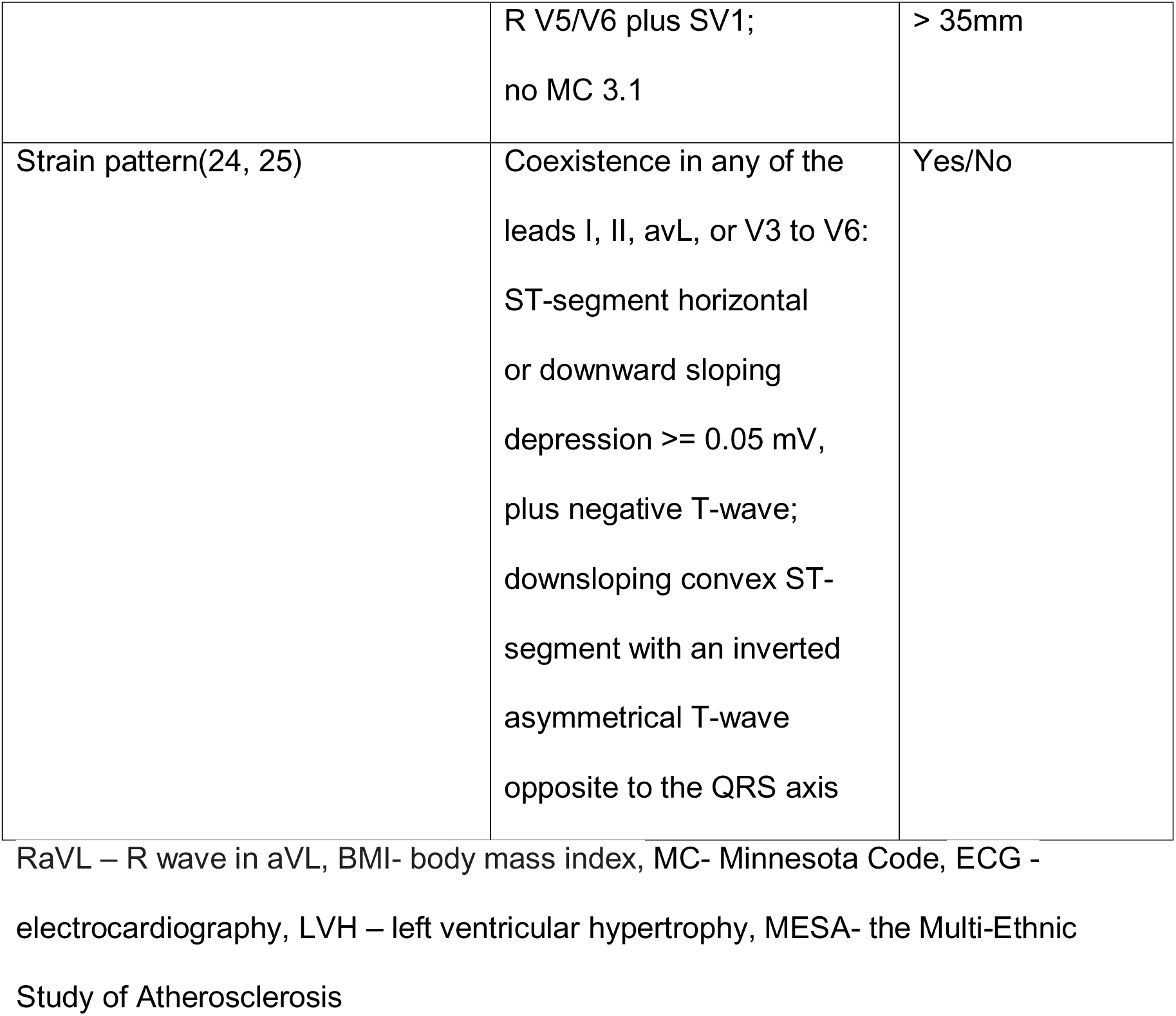
Characteristics of electrocardiographic indexes used in this study.

#### Focused transthoracic echocardiography

Focused echocardiography (fTTE) was performed at baseline by trained study personnel using standardized acquisition protocols and centrally reviewed under the supervision of board certified cardiologists supported by an automated FDA-approved deep-learning workflow (Us2.ai, Singapore) (26, 27). LVM, LVM indexed by BSA and relative wall thickness (RWT) were calculated according to guideline recommendations (LVM defined as 0.8x 1.04x [(IVS + LVID + PWT)^3-LVID^3] + 0.6 grams, where IVS is interventricular septum diastole (unit mm), LVID is left ventricular end-diastolic diameter (units mm), and PWT is posterior wall diastole (units mm)(28). LVH was defined as LVMI >95g/m^2^ in women, >115g/m^2^ in men (2).

### Statistics

For testing the normality of distribution, a Shapiro-Wilk test was used. Normal distribution was defined as a p-value ≥0.05. Continuous data were reported as median (interquartile range), categorical data as n (% of total). Differences between groups were calculated using a Wilcoxon signed rank test for continuous data and using a chi-square test for categorical data.

#### Testing of ECG parameters

Correlations were calculated with the spearman method using the ggpubr-package (29). Correlations were ranked from (0.01-0.19) indicating no or negligible relationship, to (≥ 0.70) indicating a very strong relationship (Table S1)(30). For accuracy analyses, specificity as (true negative) / (true negative + false positive), sensitivity as (true positive) / (true positive + false negative) was calculated. The negative predictive value (NPV) was calculated as (true negative) / (true negative + false negative). The positive predictive value (PPV) was calculated as (true positive) / (true positive + false positive). The accuracy was calculated as (true positive + true negative) / N. Receiver operating characteristics (ROC) curves were plotted and area under the curves (AUCs) were calculated using the pROC package(31),(32). Discriminative property was graded: AUC of 0.5 suggested no discrimination, 0.7 to 0.8 was considered acceptable, 0.8 to 0.9 was considered excellent, and more than 0.9 was considered outstanding(32). All calculations were completed by using R (33).

#### Selection of the best ECG-parameters to improve cut-off values

The four best parameters were then selected according to their AUC for further evaluation regarding improved cut-off values for this cohort. Furthermore, the general MESA ECG-LVH index, as the best ECG-LVH parameter for African Americans in the Mesa Study was used(10). For each of the parameters test characteristics were calculated for the standard threshold, 90% and 95% accuracy, 90% and 95% sensitivity, 90% and 95% specificity and the highest Youden index (J), being defined as the point in the AUC curve with the farthest distance to the diagonal line; calculated as highest value (J) =sensitivity+specificity−1. Discriminative performance for the new cutoffs of each of the parameters were calculated using the pROC package (31).

## RESULTS

### Characteristics of the Patient Cohort

Of 1 268 CoArtHA participants, 1 125 (49% from Tanzania, 51% from Lesotho) had evaluable ECG and echocardiography data (Figure 2). Median age was 53.4 (IQR:44.6-64.4) years, 28% were male, and 38% were HIV positive. LVH by echocardiography was identified in 56 participants (5%). Participants with LVH were older, more often female, more often with a history of hypertension and had higher systolic and diastolic blood pressure, and body mass index compared with those without LVH (Table 2).

**Table 2:** Baseline demographic and clinical characteristics of the study sample, overall and regarding subgroups with echocardiographic signs for LVH.

| Variables | Overall<br>(n=1125) | fTTE- LVH<br>present<br>(n=56) | fTTE- LVH<br>absent<br>(n=1069) | p-value |
| --- | --- | --- | --- | --- |
| Tanzania, n (%) | 553 (49.2) | 47 (83.9) | 506 (47.3) | < 0.001 |
| Lesotho, n (%) | 572 (50.8) | 9 (16.1) | 563 (52.7) | <0.001 |
| Age, years,<br>median (IQR) | 53.4 (44.6 –<br>64.4) | 59.0 (49.1 –<br>68.1) | 53.2 (44.3 –<br>64.3) | 0.009 |
| Male, n (%) | 314 (27.9) | 8 (14.3) | 306 (28.6) | 0.029 |
| HIV-positive, n<br>(%) | 423 (37.6) | 10 (17.9) | 413 (38.6) | 0.003 |
| Previously<br>diagnosed HT, n<br>(%) | 469 (41.7) | 34 (60.7) | 435 (40.7) | 0.012 |
| Diabetes, n (%) | 22 (2.0) | 0 | 22 (2.1) | 0.323 |
| Diabetes never<br>checked, n (%) | 102 (9.1) | 3 (5.4) | 99 (9.3) |  |
| BMI, (kg/m<sup>2</sup>),<br>median (IQR) | 26.4 (22.8 –<br>30.7) | 25.1 (21.5 –<br>28.1) | 26.5 (22.8 –<br>30.7) | 0.032 |
| BSA, (m<sup>2</sup>),<br>median (IQR) | 1.71 (1.55 –<br>1.85) | 1.57 (1.47 –<br>1.76) | 1.70 (1.56 –<br>1.85) | 0.001 |
| Systolic BP<br>(mmHg), median<br>(IQR) | 149 (141 –<br>163) | 165 (152 –<br>182) | 148 (140 –<br>161) | <0.001 |
| Diastolic BP<br>(mmHg), median<br>(IQR) | 99 (93 – 106) | 104 (98 –<br>116) | 98 (93 – 105) | <0.001 |

fTTE - focused echocardiography, HT – hypertension, LVH-left ventricular
hypertrophy, BMI - body mass index, BP- blood pressure, BSA- body surface area

### Prevalence of LVH ECG markers

The overall prevalence of positive ECG markers according to the presence of echocardiographic signs for LVH is shown in Table 3. The highest prevalence of positive ECG LVH markers was observed with the general MESA ECG-LVH criterion (633/1125 [56.3%]) participants), the ethnicity- and gender-specific MESA ECG-LVH criterion (566/1125 [50.3%]) participants) and Sokolow-Lyon voltage index (390/1125 [34.7%]) participants. There were no significant differences in the prevalence of positive values of these criteria, as well as for the Framingham-adjusted Cornell voltage index, and Minnesota Code 3.3, between the group of patients fTTE - LVH and the patients without fTTE-LVH (p>0.05).

**Table 3.**
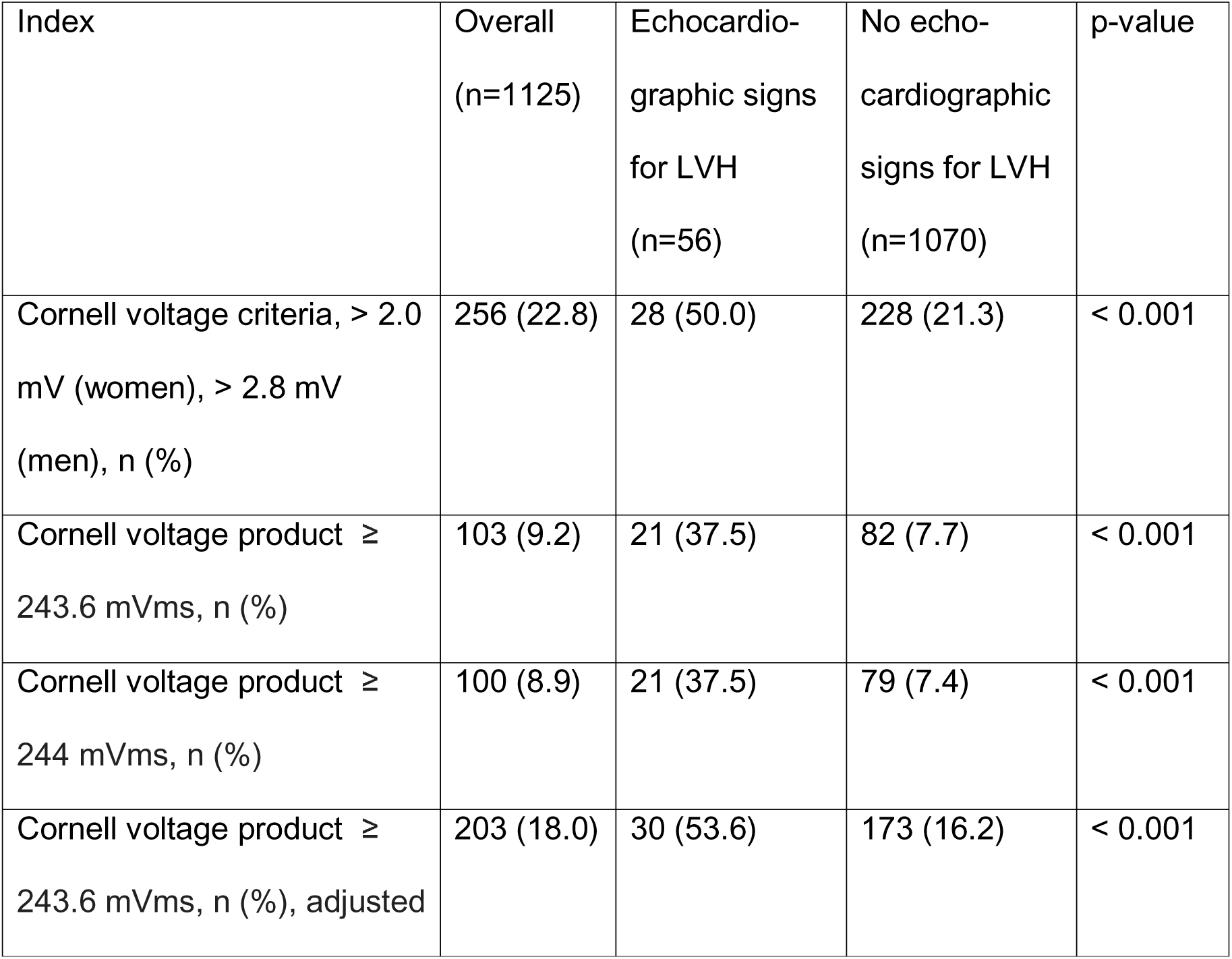
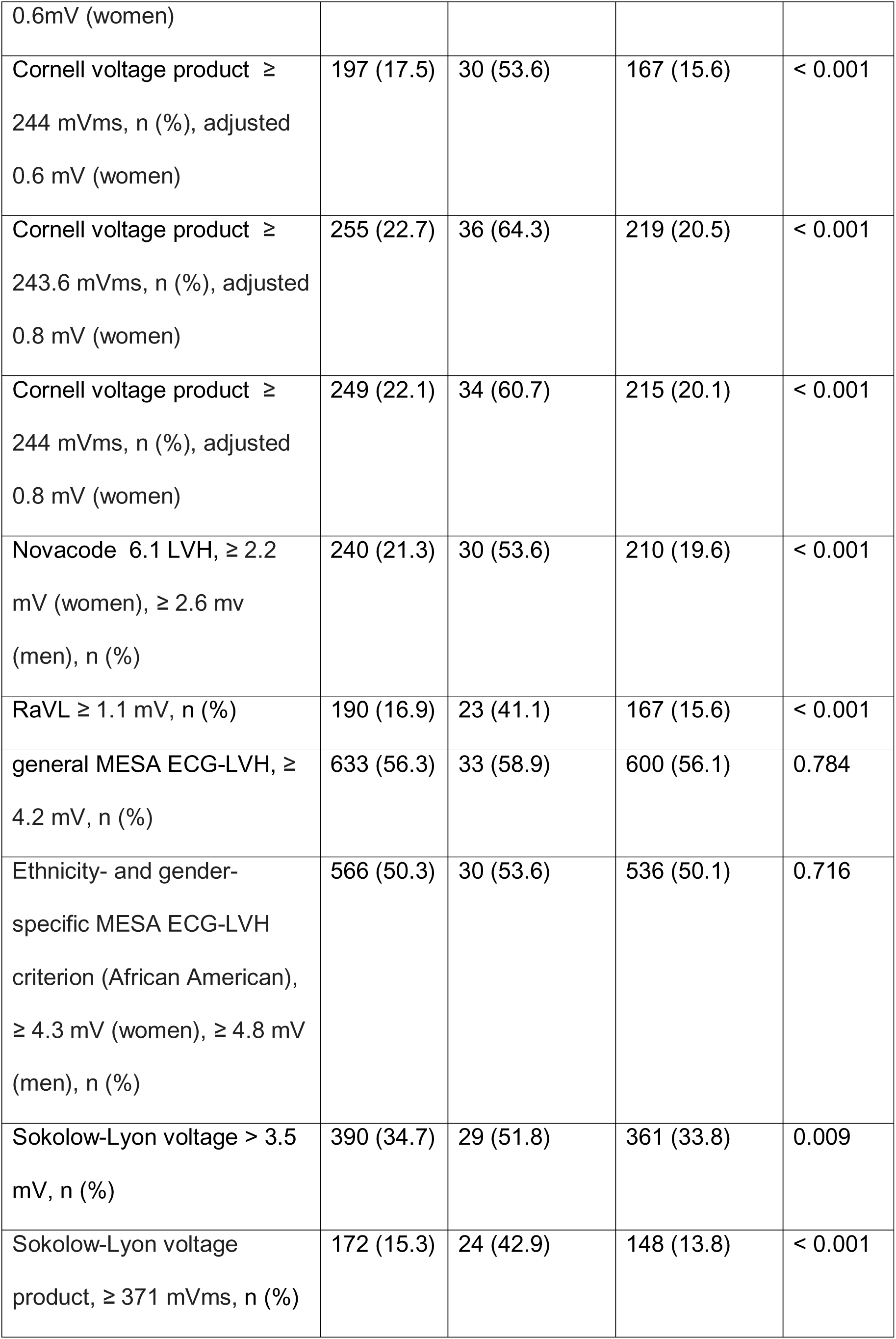
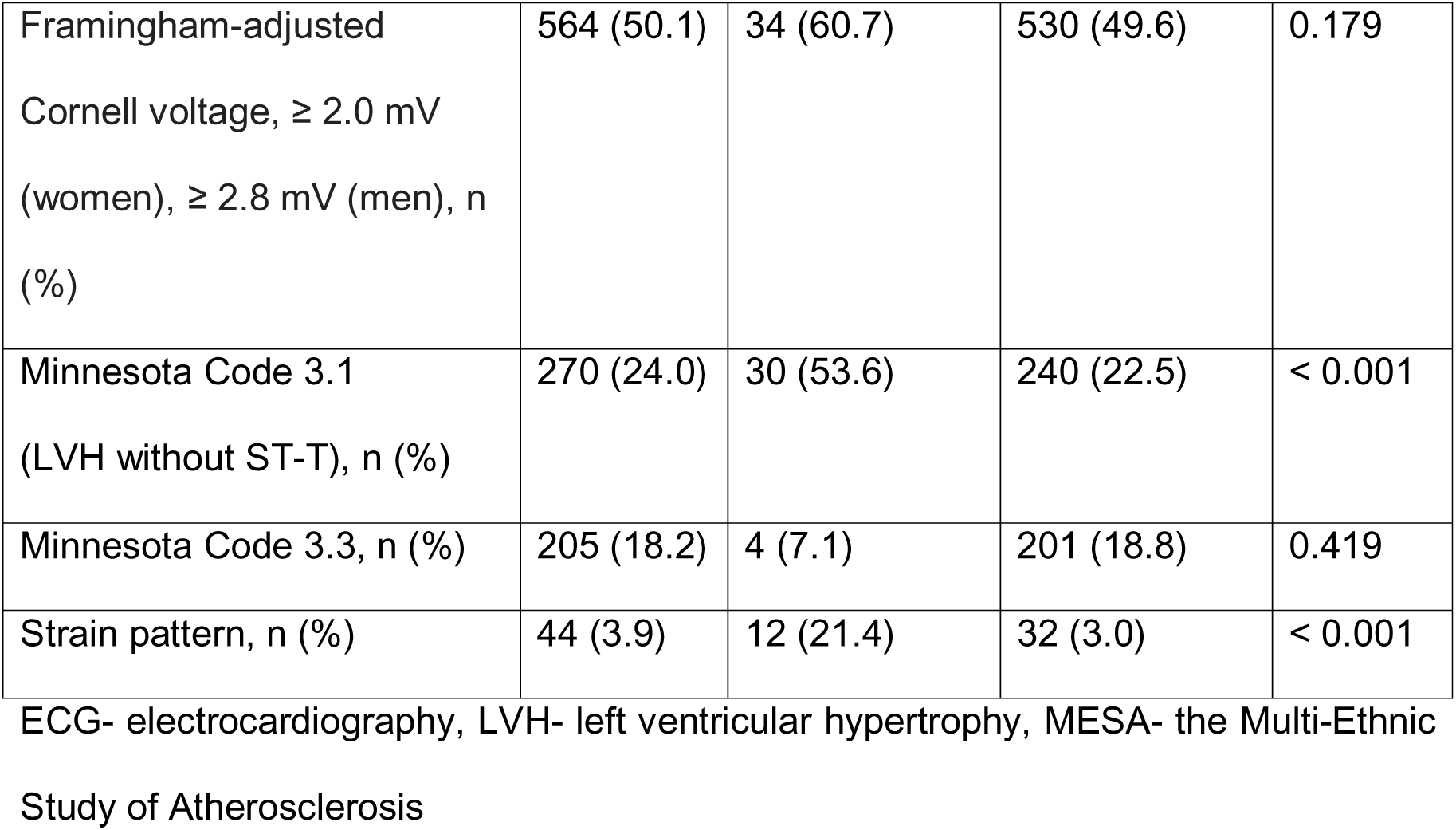
Prevalence of positive ECG markers in the entire cohort, as well as separated for the group with echocardiographic signs of LVH and those without.

### Characteristics of ECG parameters

Median values of the continuous ECG criteria in the group of participants with fTTE-LVH were significantly higher than in the group without fTTE-LVH, except for the General and male specific MESA ECG-LVH criterion and the male and female specific Framingham-adjusted Cornell voltage (Table S2).

### Correlations between LVMI and ECG-LVH criteria

Continuous LVH ECG criteria were compared with continuous LVMI to determine the degree of correlation between these methods and to evaluate the extent to which ECG parameters can serve as markers for echocardiographic changes (Figure S1). Significant, weak to moderate, positive correlations were found between continuous LVMI and all ECG indices, with the highest correlations observed for unadjusted Cornell voltage product, Cornell voltage criteria, 0.6mV adjusted Cornell voltage product and R amplitude aVL (rho = 0.373, rho = 0.327, rho = 0.324 and rho = 0.304, respectively; p < 0.001 for all). The MESA ECG-LVH criteria showed a significant positive but very weak correlation with LVMI (rho = 0.179, p < 0.001).

### Diagnostic performance of ECG-LVH criteria

Sensitivity, specificity, positive (PPV) and negative (NPV) predictive values and the AUC for LVH were calculated for each ECG criterion with recommended standard cut-off values in comparison to echocardiographic signs of LVH (Table 4).

**Table 4:**
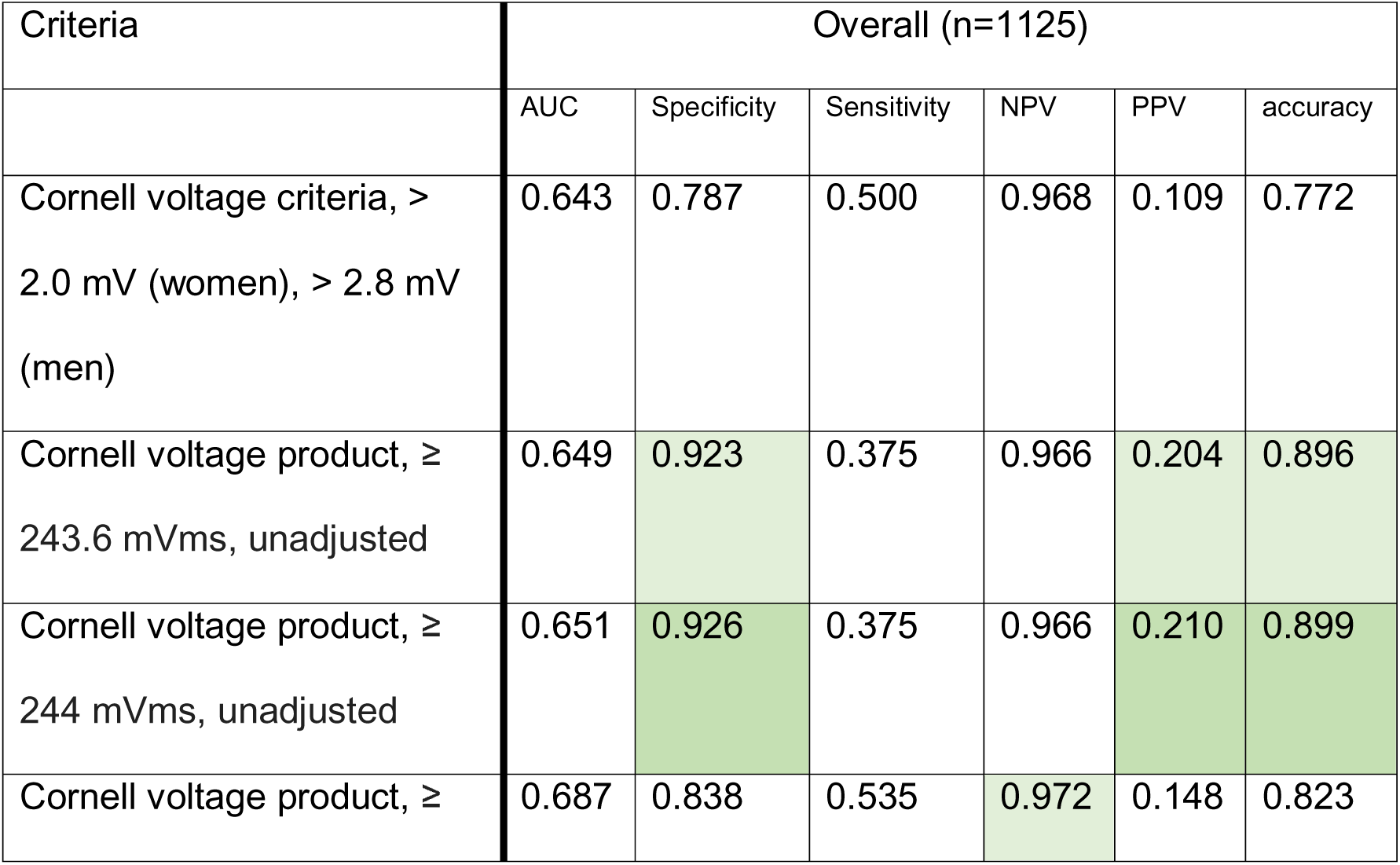
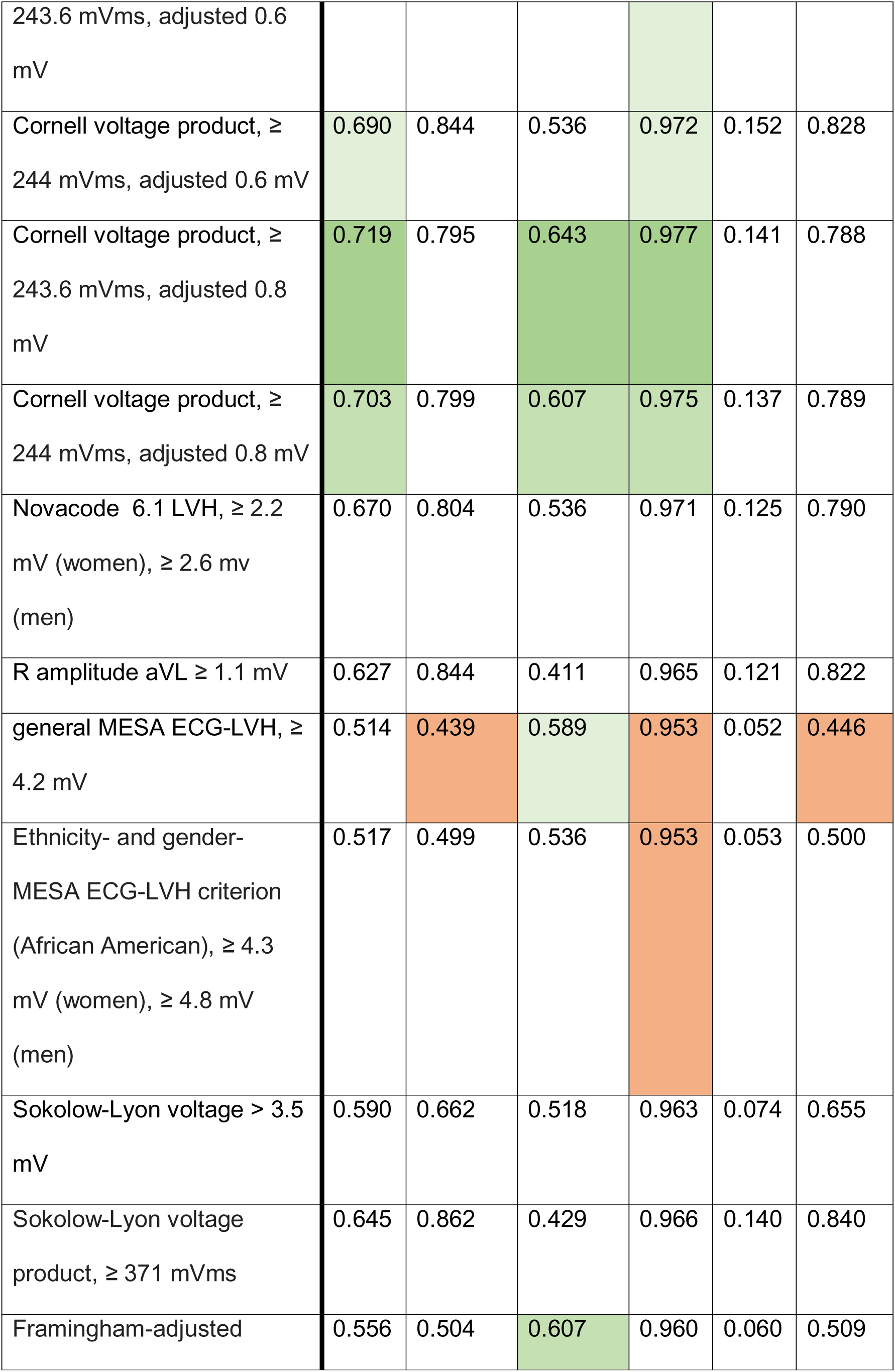
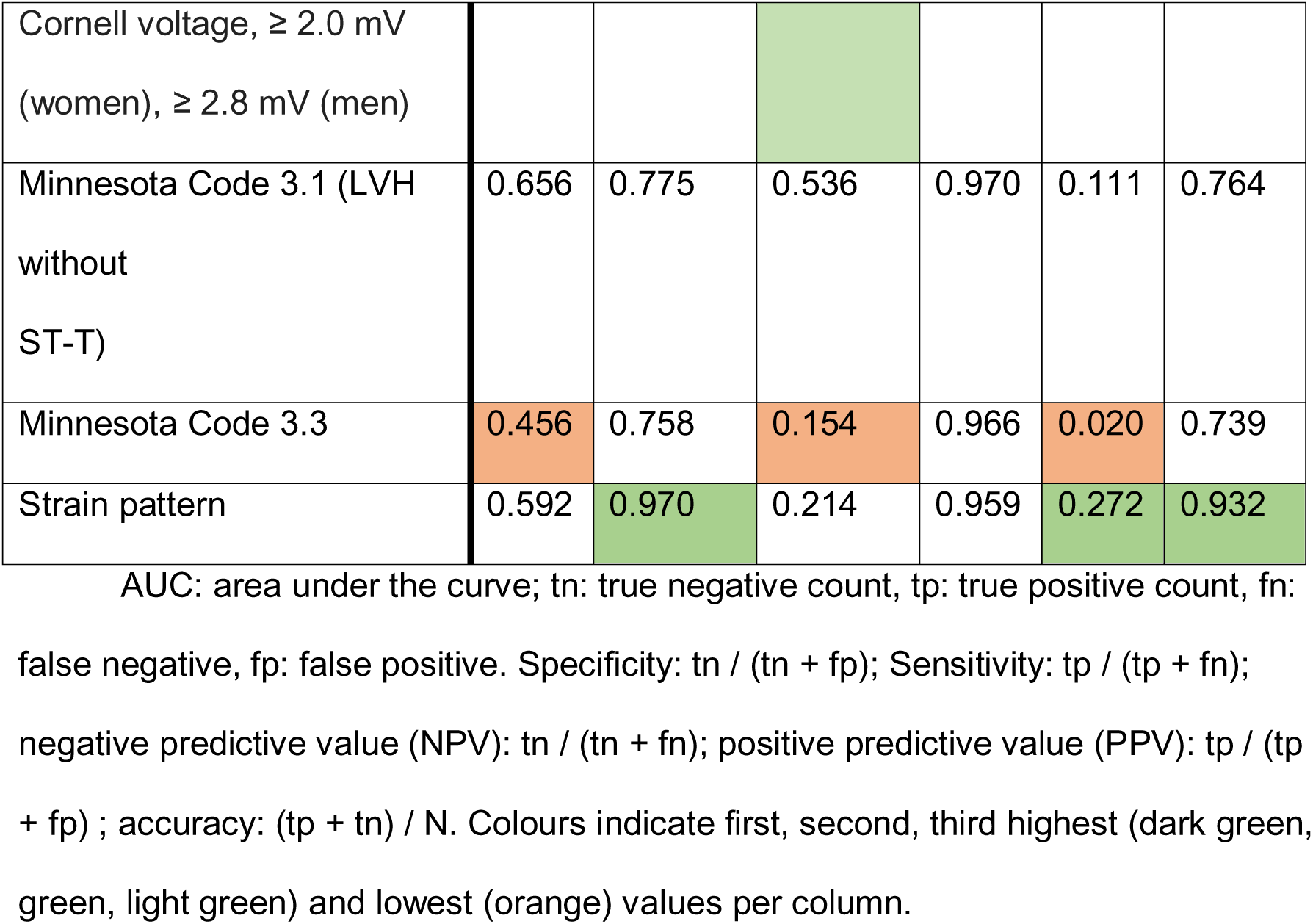
Performance of standard ECG LVH criteria in overall study population.

Using standard cut-offs, the Cornell voltage product (adjusted + 0.8 mV for women) achieved the highest AUC (0.719) with sensitivity 64% and specificity 80%, followed by the 0.6 mV-adjusted (AUC 0.690) and unadjusted (AUC 0.651) versions. Strain pattern and unadjusted Cornell voltage product yielded the highest specificity (> 90%) but lower sensitivity. MESA-derived criteria showed poor discrimination (AUC 0.517).

### Analysis of continuous ECG parameters to identify most promising ECG parameter as an indicator for LVH Detection

The ROC plot in Figure 3 provides a comparative analysis of the diagnostic performance of the studied ECG criteria as continuous values with regard to the presence of LVH to see if an improvement of the cut-off values would be possible. Among the indexes investigated, compared to the ‘neutral’ AUC value of 0.5, the Cornell voltage product adjusted with 0.8mV and 0.6mV (women), the unadjusted Cornell voltage product, and the R wave amplitude in aVL had the highest AUC values (0.765, 0.761, 0.738, and 0.708, respectively) with an acceptable discriminative property (32). The MESA ECG-LVH index showed the lowest AUC of the tested parameters (0.577).

**Figure 3.**
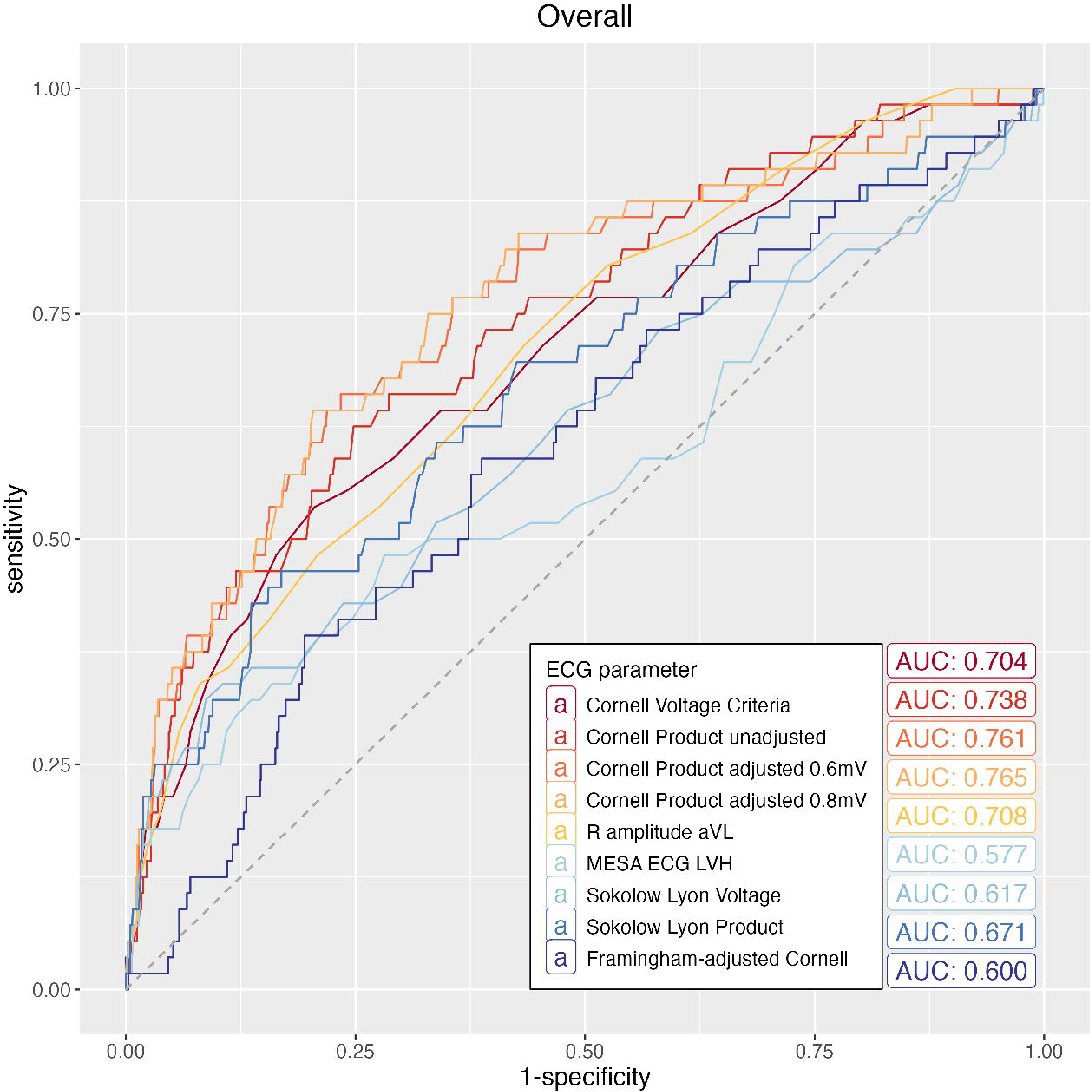
ROC plot with area under the curve (AUC) for continuous electrocardiography (ECG) parameters. Legend: ECG-electrocardiography, LVH-left ventricular hypertrophy, MESA-the Multi-Ethnic Study of Atherosclerosis.

### Alternative cut off values for selected ECG parameters

Based on the previous results, we selected the three best criteria for further evaluation: Cornell voltage criteria, Cornell voltage product, and R amplitude aVL, as well as the MESA ECG-LVH index. We analysed both standard and optimized thresholds for these ECG LVH criteria to identify the most clinically applicable thresholds (Table S3, Figure S2). Considering the limited number of men diagnosed with fTTE-LVH in our cohort, we evaluated novel thresholds for the Cornell voltage criteria without sex-specific adjustments. Figure S2 illustrates the discriminative performance of each parameter using both traditional cut-off values and thresholds optimized for maximal Youden’s index. It also reports the AUCs corresponding to cut-offs that achieve sensitivity, specificity, or accuracy greater than 90% and 95%, respectively (Table S3). Highest AUC (0.719) was calculated for the traditional cut-off of the Cornell voltage product adjusted with 0.8 mV (women). Figure 4 summarizes the best test and cut-off values for highest Youden index/AUC, sensitivity >90% and specificity >90% for the Cornell voltage product adjusted with 0.8 mV (women), despite AUC for sensitivity >90% was numerically higher for the unadjusted Cornell voltage product (0.599 vs. 0.620).

**Figure 4.**
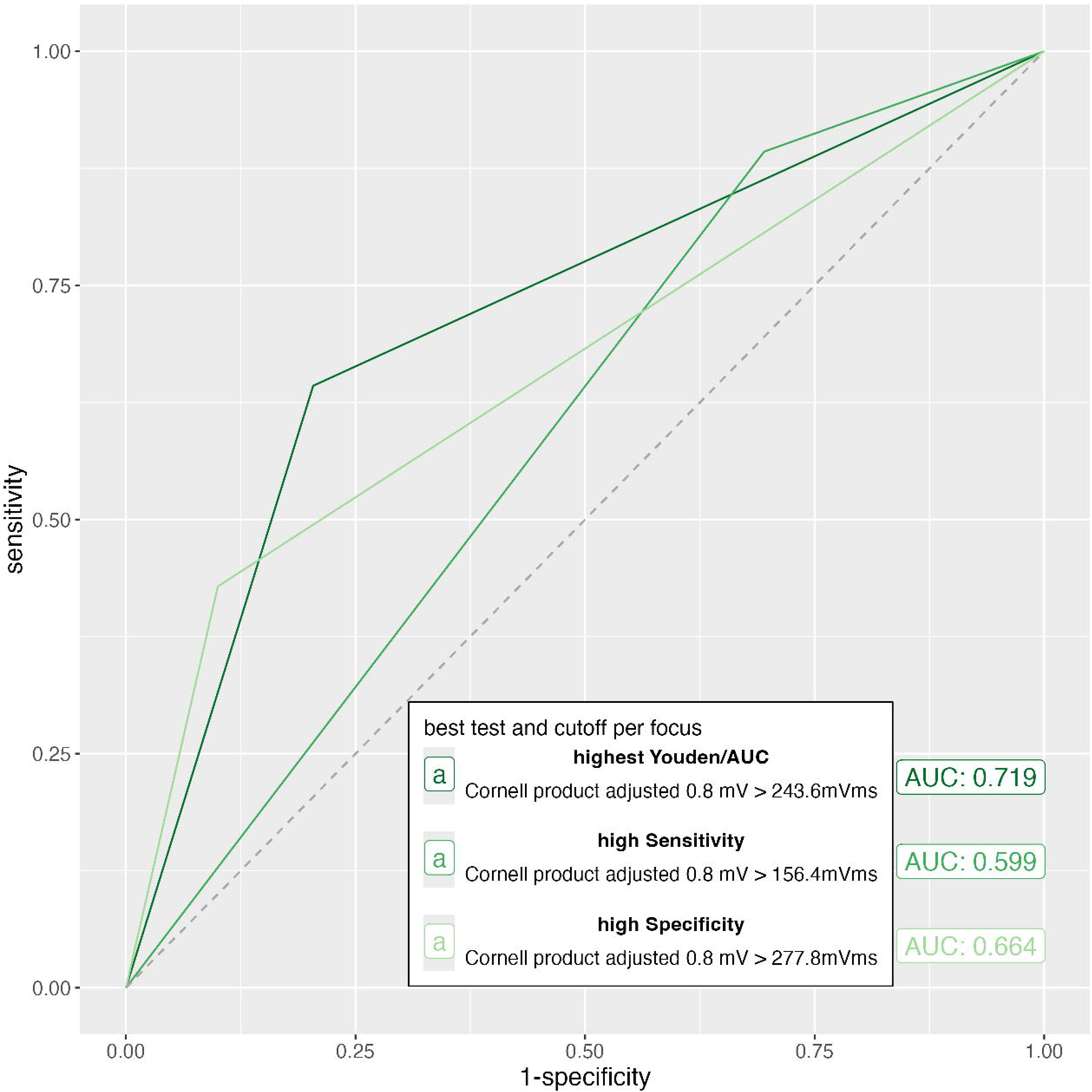
Discriminative performance of the Cornell voltage product adjusted with 0.8 mV (women) at the highest Youden index/AUC derived cut-offs and sensitivity >90% and specificity >90% cut-offs.

## DISCUSSION

This sub-analysis of the CoArtHA trial provides several important insights into the diagnostic utility of ECG criteria for left ventricular hypertrophy (LVH) among hypertensive Black African adults.

First, LVH ECG indices showed a weak to moderate correlation with echocardiographic LV mass index (LVMI), with the Cornell voltage product and its female-adjusted version performing best. Second, appliying traditional cut-off values yielded highest discriminative properties for the Cornell voltage product with 0.8 mV adjustment for women at a cut-off of 243.6 mVms, followed by the cuf-off of 244 mVms. Third, the limited sensitivity of ECG in diagnosing LVH highlights the need for tests with higher sensitivity and acceptable specificity. Rodrigues et al e.g. proposed lowering sex-specific thresholds for the Cornell voltage criteria to improve sensitivity (34). The CoArtHa study also aimed to enhance ECG criteria for LVH in African populations, supporting ethnic-specific thresholds. The Youden index is commonly used to identify optimal cut-off values, minimizing both false positives and negatives (35). Within our cohort novel cut-off values only have better discriminative properties than standard cut-offs, when high sensitivities or specifities are aimed for. Interestingly, the highest AUC for the detection of LVH was reached at the best Youden Index with the Cornell voltage product with adjustment of 0.8 mV for women at the cut-off of 243.6 mVms, showing a sensitivity of 64.3% and specificity of 79.6%. ECG criteria for LVH were originally designed to reach a specificity as high as 90% to 95%, which in our cohort was achieved only by the unadjusted Cornell voltage product and Strain pattern at standard cut-offs, with the latter having a lower sensitivity of 21% compared to 37.5% (36). An adapted best threshold for > 90% specificity could be using the Cornell voltage product adjusted with 0.8 mV for women at >277 mVms with a sensitivity of 42.9%. Regarding Cornell voltage criteria best Youden index at 2.25 mV showed higher AUC values and improved sensitivity to 53.6%, while maintaining specificity at a level of 79.4%. Further studies are needed to confirm the clinical usefulness of these thresholds for diagnosing LVH in Black African hypertensive patients.

Collectively, these findings demonstrate that standard ECG criteria especially the Cornell voltage product with a 0.8 mV adjustment for women remain applicable but may require minor threshold adjustments when used in African populations with untreated hypertension.

### Several aspects of the present study deserve to be mentioned

The weak to moderate correlations between ECG-LVH indices and echocardiographic LV mass index observed in our study are consistent with prior reports from both African and non-African cohorts.(5, 6, 9, 34, 36–38)

Our findings of variable but overall low-to-moderate sensitivity (0.15–0.59) combined with higher specificity (0.44–0.97) are consistent with previous studies in African adults with hypertension, which have repeatedly reported that standard ECG criteria for LVH show limited sensitivity but good specificity (5, 9, 36, 39, 40). In a study conducted in Nigeria in 90 subjects evaluating four ECG criteria, Cornell criterion and Sokolow-Lyon criterion showed a lower accuracy than our results, but the defined cut off for LVH in TTE was >125/m^2^ for both sexes which is higher than the recommended gender-specific cutoff values we used (9). In a subgroup of 120 African American participants of the LIFE study, Sokolow-Lyon overestimated and Cornell voltage criteria underestimated the presence of LVH relative to white participants compared to TTE, in terms of sensitivity and specificity – but not in terms of overall test performance in ROC curve analysis (41). In line with our results, Cornell voltage product had the highest numerical AUC value of 0.665 in the LIFE study (41). Interestingly and in contrast to a study by Jain et al, comparing ECG parameter to cardiac MRI in several ethnicities within the MESA study and deriving the MESA ECG-LVH criteria, the general MESA ECG-LVH and the ethnicity- and gender-specific MESA ECG-LVH criterion had a low specificity and sensitivity in our cohort.(10) Within the MESA cohort and when applying ethnicity and gender specific cut-offs, specificity and sensitivity for the participants of African American descent was 90% and 37-39%, respectively with an ROC AUC of 0.64.In addition to the geographical differences of the trials namely United States versus sub saharan Africa, there were also differences between the cohorts with a 50% proportion of males and a higher prevalence of MRI-LVH of 11% in the MESA study. Additionally participants were older and had an higher BMI. Another reason for the discrepancy may be the differences between MRI and TTE, but again in the MESA study, the agreement for LVH between the imaging modalities ranged from 77-98% with a kappa coefficient from 0.1-0.76(42). To our knowledge, the MESA criteria had not been externally validated.

Generally, in the CoArtHa study three particular ECG criteria (Cornell voltage сriteria, Cornell voltage product (especially with adjustment), and R amplitude aVL) had better discriminative properties and were more sensitive than other criteria. This aligns with other studies in different ethnic populations that demonstrated that the Cornell-based criteria had superior diagnostic performance compared to Sokolov-Lyon, and other criteria (34, 40, 43, 44).

Regarding the prognostic value, presence of LVH is an important marker for mortality, especially in African American patients, making screening of this condition even more relevant(41, 45, 46). This is true not only for LVH diagnosed by TTE or MRI but also for LVH detected by ECG criteria in different settings, even though the prevalence of LVH by ECG is higher than by imaging.

For example, within the LIFE study, regression of LVH defined by Cornell voltage criteria, Sokolov-Lyon voltage criteria or ECG Strain pattern was associated with an improved cardiovascular outcome independent of BP reduction (47). Additionally, presence of these markers at baseline or development of new Strain pattern over time were predictors of cardiovascular events (47). Moreover, in a meta-analysis conducted by Zhigang You et al, Cornell voltage criteria had a stronger predictive value for cardiovascular risk than the Sokolov-Lyon index and Cornell voltage product in general populations, without studies coming from the African continent. (48).

Of note, there is evidence that the information from TTE and ECG may complement each other, with, however, a higher impact of the TTE diagnosis. A study by Seko and colleagues evaluated the prognostic relevance of presence and absence of LVH in TTE and ECG by using Cornell product in hospitalized asian participants. When considering the resulting four possible patterns of TTE/ECG, namely +/+, +/-, -/+ and -/-, the hazard ratio for major adverse cardiovascular events over 3 years was 1.63 (1.16-2.28), 1.68 (1.23-2.30) and 1.09 (0.85-1.41) compared to patients no evidence of LVH(49). Therefore, ECG remains a useful tool for risk stratification in hypertension not only in countries where poor technical provision and costs may limit the use of echocardiography. Therefore, it may be clinically feasible to use ECG with tailored cutoff values to identify a group of patients at lower risk for LVH and deprioritise them for further imaging.

Our study has several limitations. Firstly, in our cohort of treatment-naïve adults with hypertension, the prevalence of fTTE LVH was 5%, resulting in 56 participants with LVH only. The low number, together with the overrepresentation of female participants within the study as well as within those with an echocardiographically confirmed LVH, makes it difficult to generalize the results, especially to men, speaking for the need of a validation in other cohorts in Africa. Secondly, the prevalence of echocardiographic LVH in hypertension varies based on LVM indexation and cut-off values. Adebiyi et al. examined different echocardiographic thresholds in 457 hypertensive native Africans, finding LVH prevalence ranged from 30.9% to 56.0%(37, 50). The highest was with LVM indexed to height²·□ (49.2 g/ht²·□ in men, 46.7 g/ht²·□ in women), and the lowest with BSA indexation (125 g/m² for both sexes)(50). This aligns with studies in southwestern Nigeria and the MAVI (MAssa Ventricolare sinistra nel soggetto Iperteso) study, which used a higher LVMI cut-off (>125 g/m²) and reported LVH prevalence of 32.2% and 27.2%, respectively(6, 9). Our analysis applies the American Society of Echocardiography definition (>115 g/m² in men, >95 g/m² in women), widely used in clinical practice and linked to adverse cardiovascular outcomes(28).

Thirdly, the fTTE in our study were not performed by fully trained cardiologists, but by doctors and nurses after a short-time training, possibly resulting in differences when compared to board certified cardiologists or cardiac sonographer. However, analyses of echocardiographies were done in a central site with the support of an artificial intelligence algorithm and interpretation of cardiologists. This approach was used also in a population-based survey by our group and has been proven to be feasible(27, 51). Additionally there is evidence that TTE image quality has limited influence on the reliability and performance of LVM assessment, when compared to cMRI(42). Finally, we could not analyse a correlation of ECG parameter with outcomes such as morbidity or mortality.

In conclusion, in this population of treatment-naïve Black Africans with uncomplicated and untreated hypertension in rural Africa, the Cornell voltage product with adjustment of 0.8 mV for women had the best diagnostic properties when compared to alternative LVH ECG criteria, with a moderate association with left ventricular myocardial mass measured by focused echocardiography, indexed to body surface area and should be the preferred ECG parameter when full or point-of-care echocardiography is not available. We found, that application of standard cut-offs are in line with best Youden Index. Specificity can be increased to 90% using a cut-off value of 277 mVms. Implementing ECG and using the best performing criteria in routine clinical care may contribute to improve the diagnosis of LVH at screening stage and may allow to identify a group of patients at lower or higher risk for hypertension-mediated cardiac damage to de-or prioritize for further echocardiography. However, given the rapid advancements in imaging technology, the increasing integration of artificial intelligence (AI) and a more comprehensive and dynamic assessment of cardiac function by fTTE, it is plausible that ECG may be gradually supplanted by fTTE in the coming years. Nonetheless, continued refinement of ECG-LVH criteria and AI-based diagnostic approaches may further enhance the utility of ECG as an accessible screening tool in resource-limited settings.

## Data Availability

The datasets used for the current study are available from the corresponding author on reasonable request.

## Acknowledgments

The authors thank all investigators, staff, and participants of the CoArtHA study for their valuable contributions.

## Supplementary Material

**Table S1.**
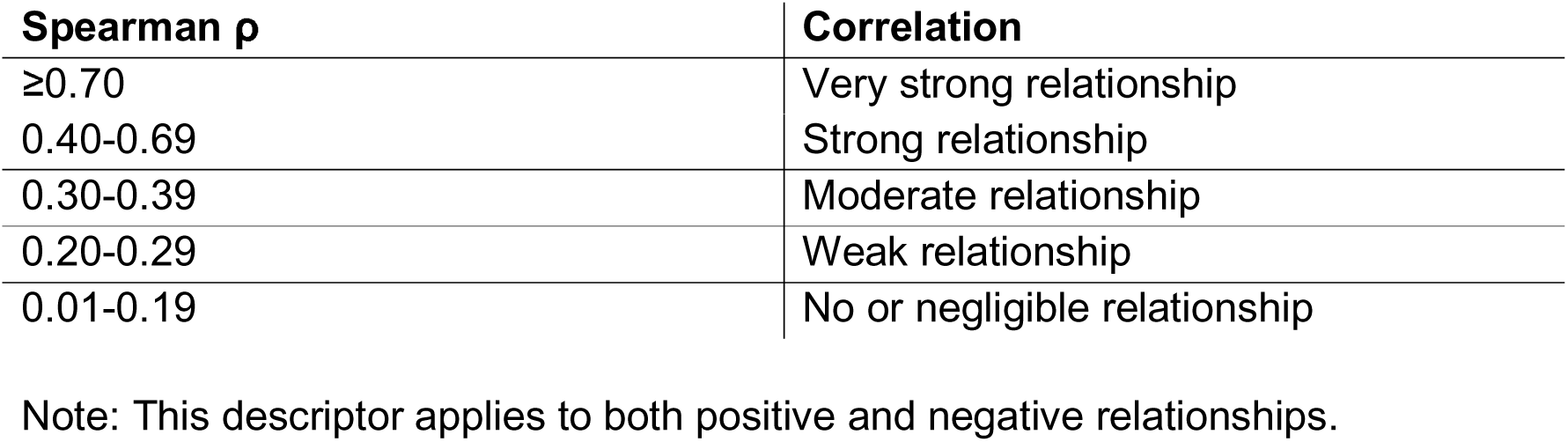
Interpretation Table of Spearman Rank-Order Correlation Coefficients (30)

**Table S2:**
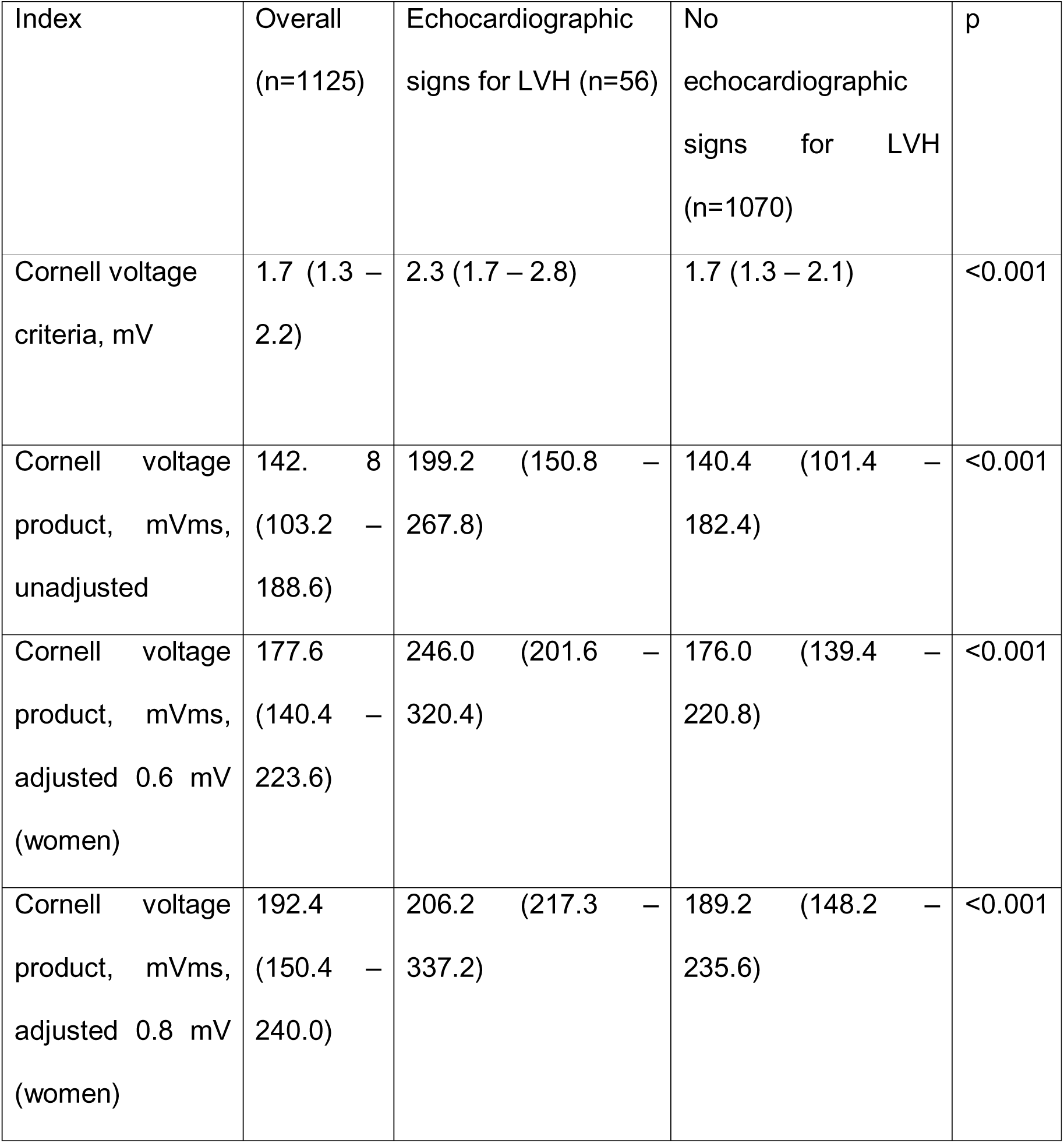
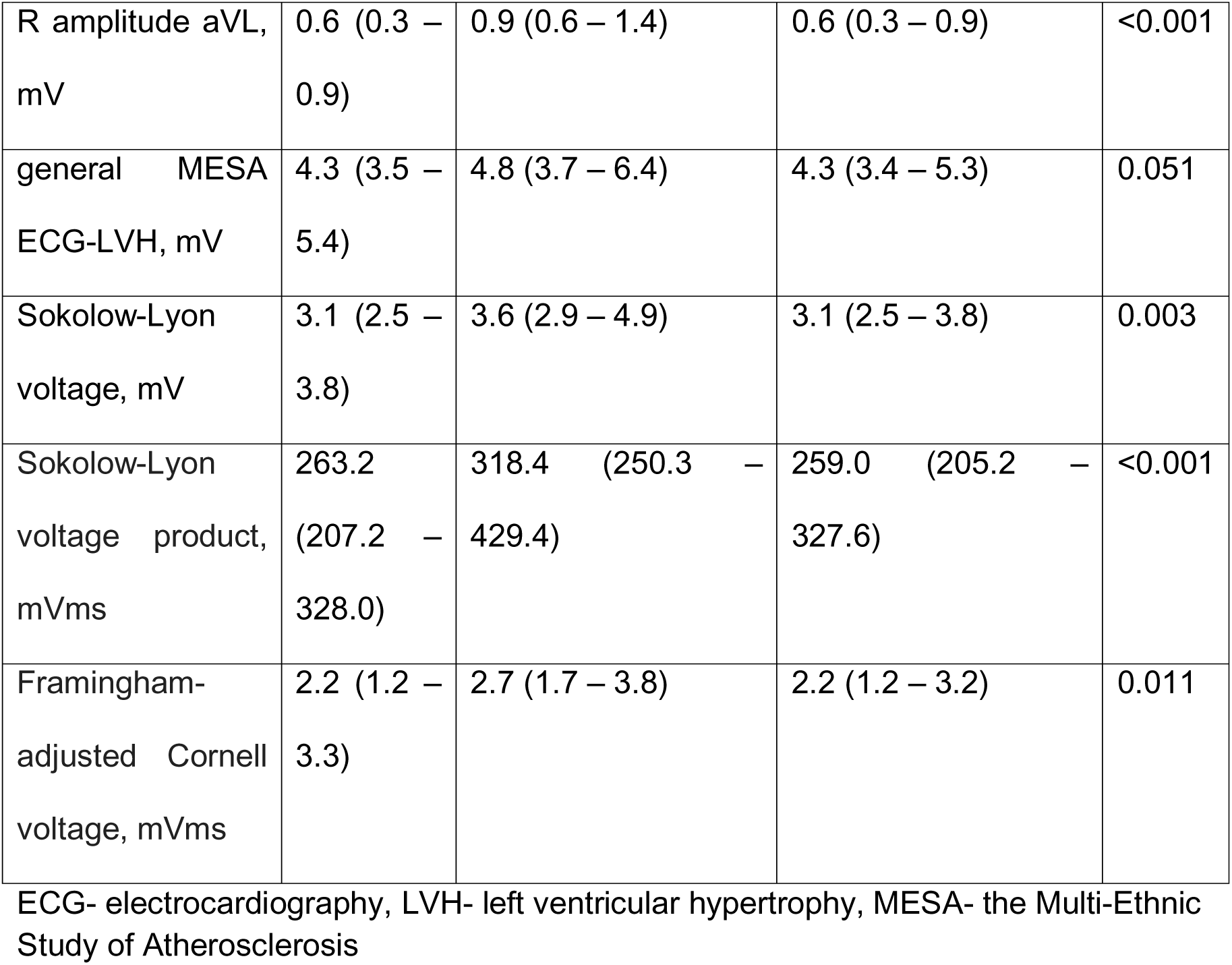
Median values (IQR) for different continuous ECG parameters overall and in participants with and without echocardiographic LVH.

| Index | Overall<br>(n=1125) | Echocardiographic<br>signs for LVH (n=56) | No<br>echocardiographic<br>signs for LVH<br>(n=1070) | p |
| --- | --- | --- | --- | --- |
| Cornell voltage<br>criteria, mV | 1.7 (1.3 – 2.2) | 2.3 (1.7 – 2.8) | 1.7 (1.3 – 2.1) | <0.001 |
| Cornell voltage<br>product, mVms,<br>unadjusted | 142.8 (103.2 – 188.6) | 199.2 (150.8 – 267.8) | 140.4 (101.4 – 182.4) | <0.001 |
| Cornell voltage<br>product, mVms,<br>adjusted 0.6 mV<br>(women) | 177.6 (140.4 – 223.6) | 246.0 (201.6 – 320.4) | 176.0 (139.4 – 220.8) | <0.001 |
| Cornell voltage<br>product, mVms,<br>adjusted 0.8 mV<br>(women) | 192.4 (150.4 – 240.0) | 206.2 (217.3 – 337.2) | 189.2 (148.2 – 235.6) | <0.001 |
| R amplitude aVL, mV | 0.6 (0.3 – 0.9) | 0.9 (0.6 – 1.4) | 0.6 (0.3 – 0.9) | <0.001 |
| general MESA ECG-LVH, mV | 4.3 (3.5 – 5.4) | 4.8 (3.7 – 6.4) | 4.3 (3.4 – 5.3) | 0.051 |
| Sokolow-Lyon voltage, mV | 3.1 (2.5 – 3.8) | 3.6 (2.9 – 4.9) | 3.1 (2.5 – 3.8) | 0.003 |
| Sokolow-Lyon voltage product, mVms | 263.2 (207.2 – 328.0) | 318.4 (250.3 – 429.4) | 259.0 (205.2 – 327.6) | <0.001 |
| Framingham-adjusted Cornell voltage, mVms | 2.2 (1.2 – 3.3) | 2.7 (1.7 – 3.8) | 2.2 (1.2 – 3.2) | 0.011 |

ECG- electrocardiography, LVH- left ventricular hypertrophy, MESA- the Multi-Ethnic Study of Atherosclerosis

**Table S3:**
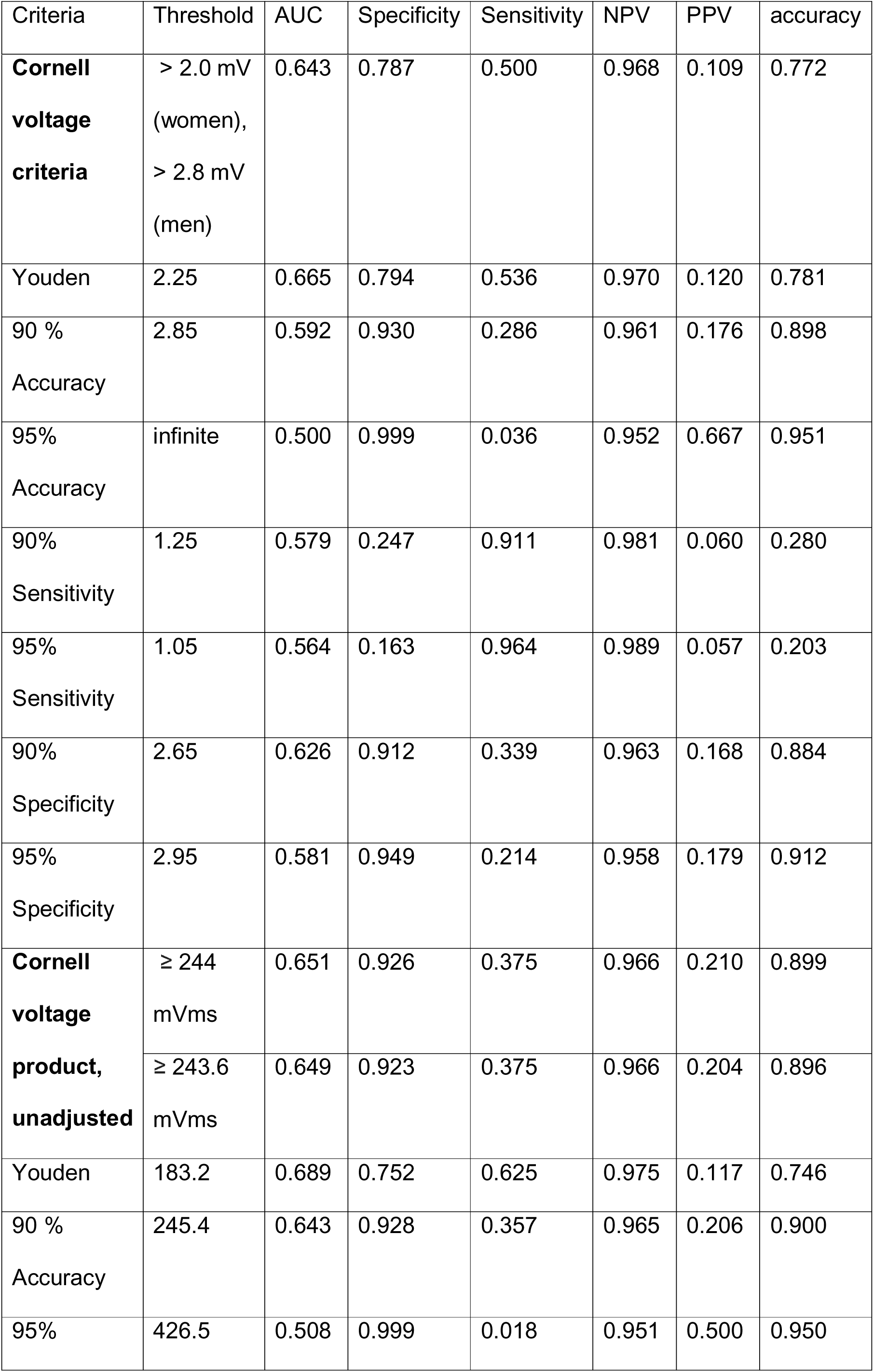
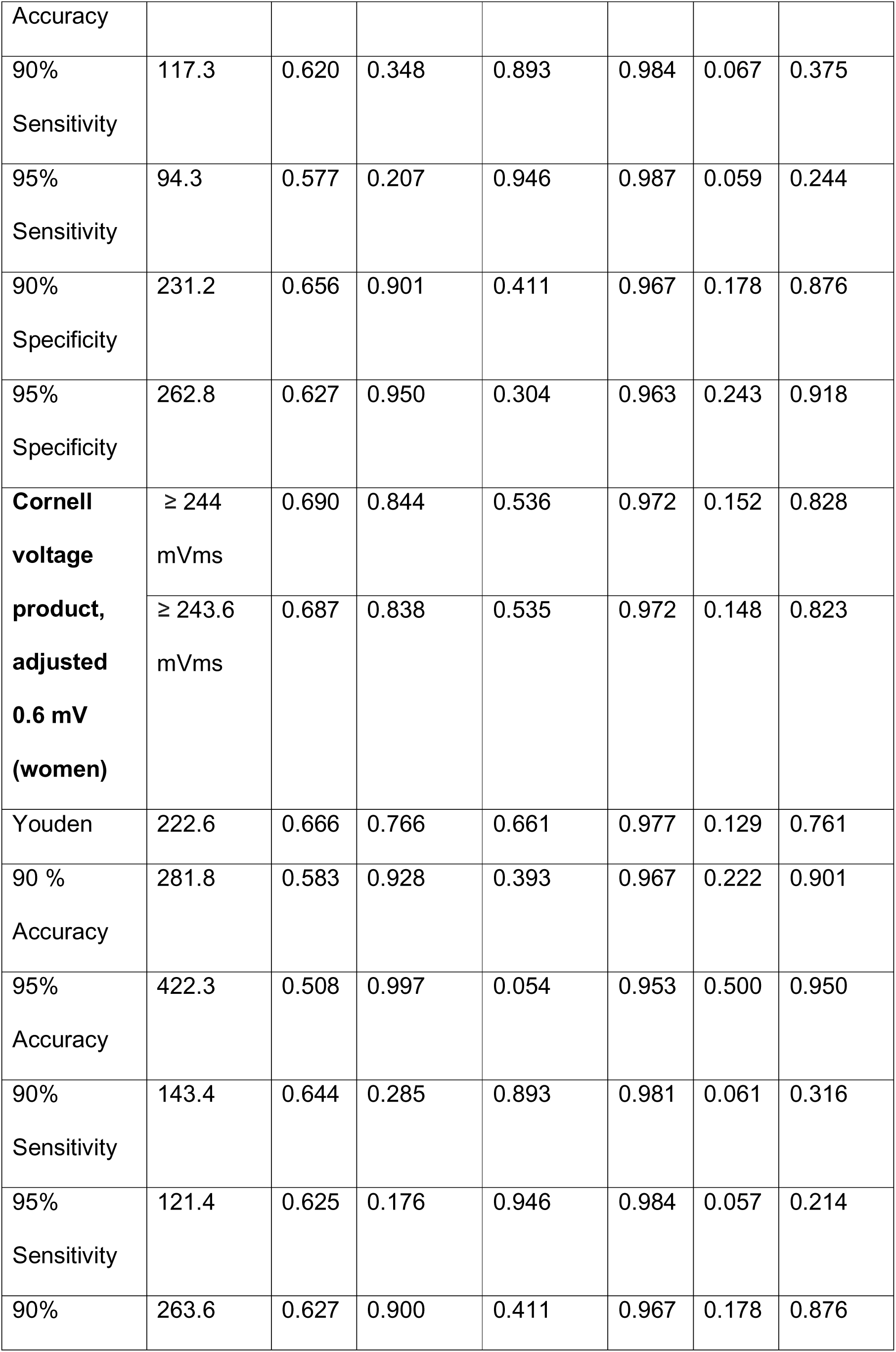
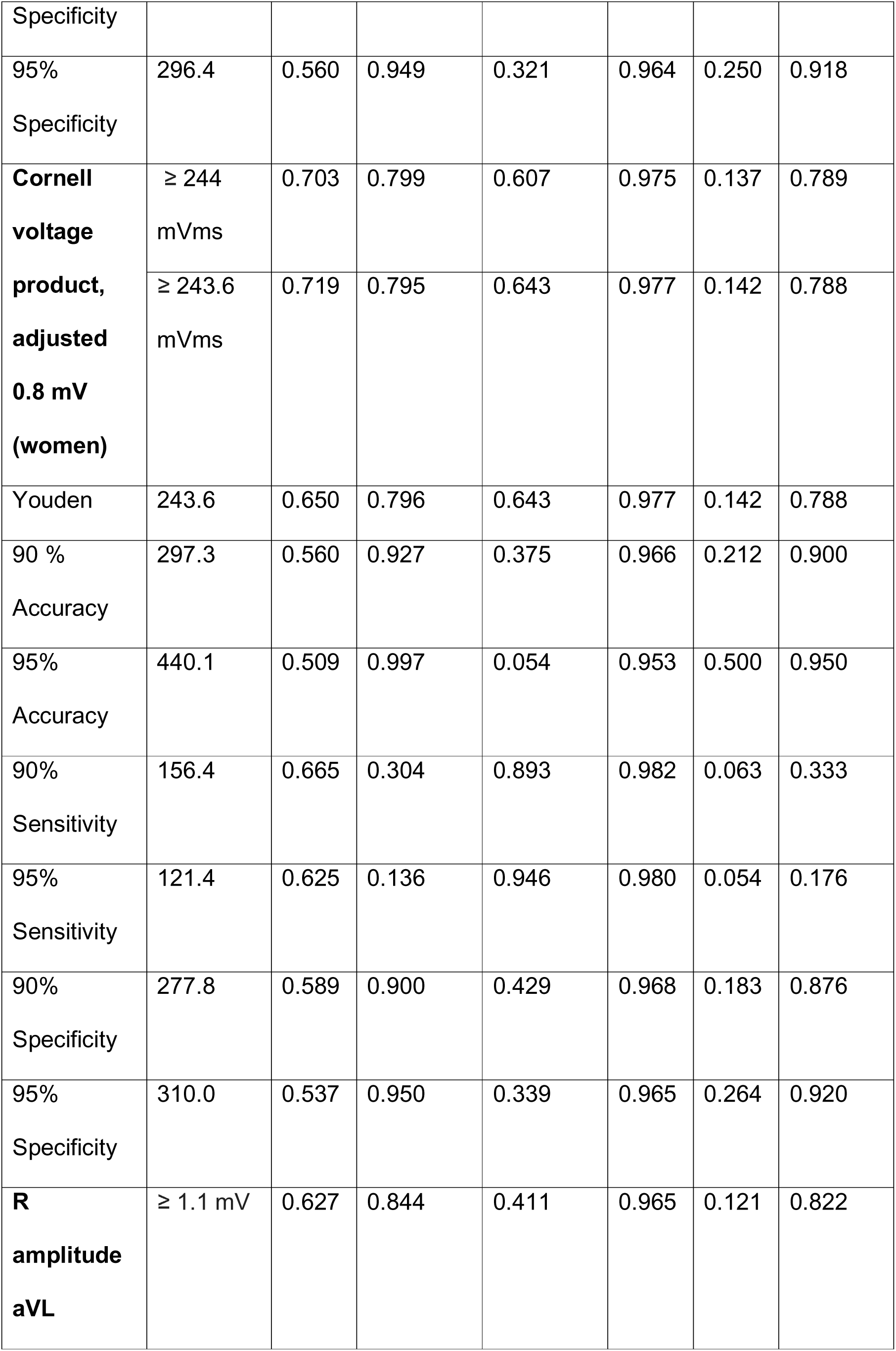
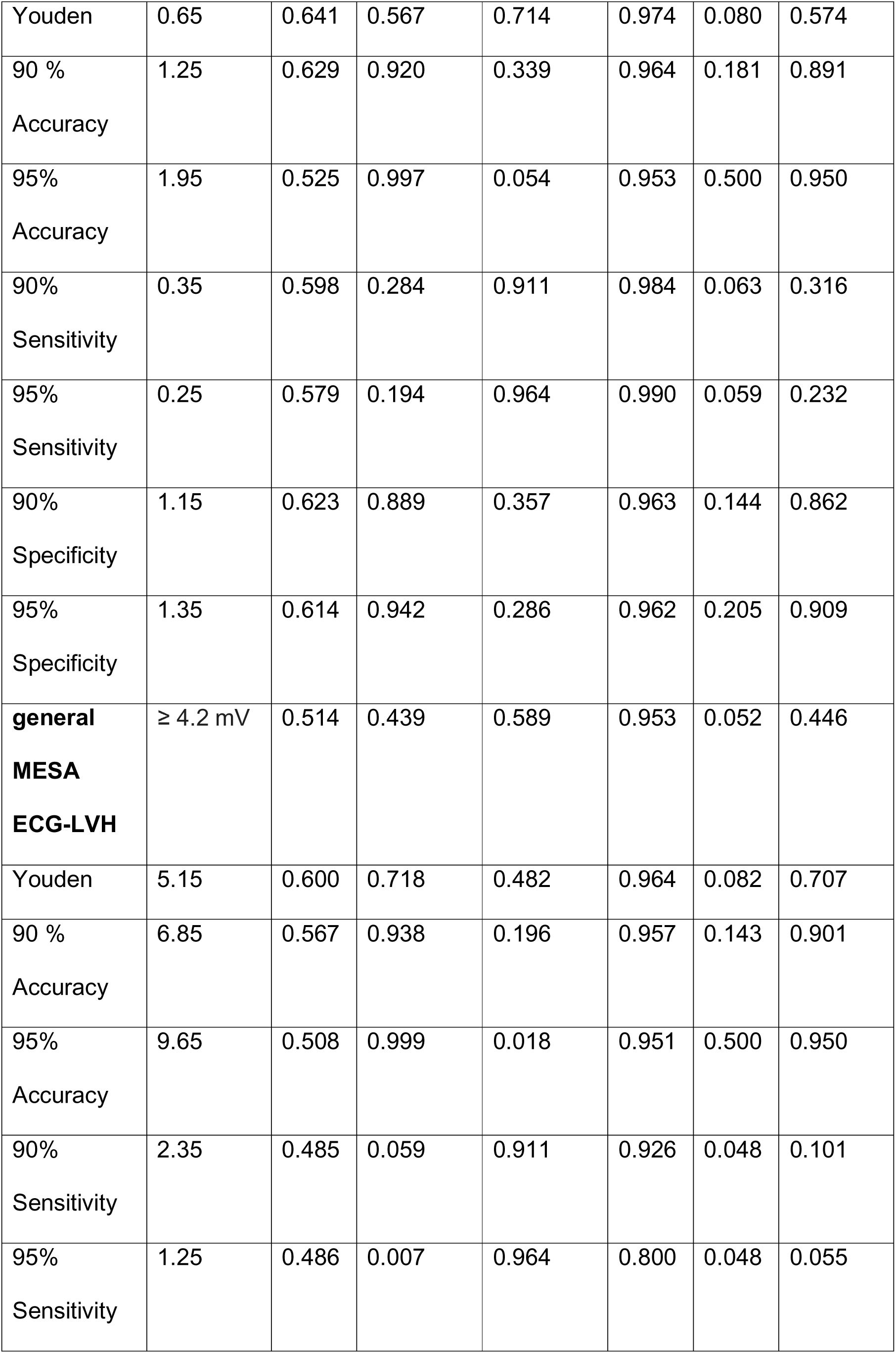
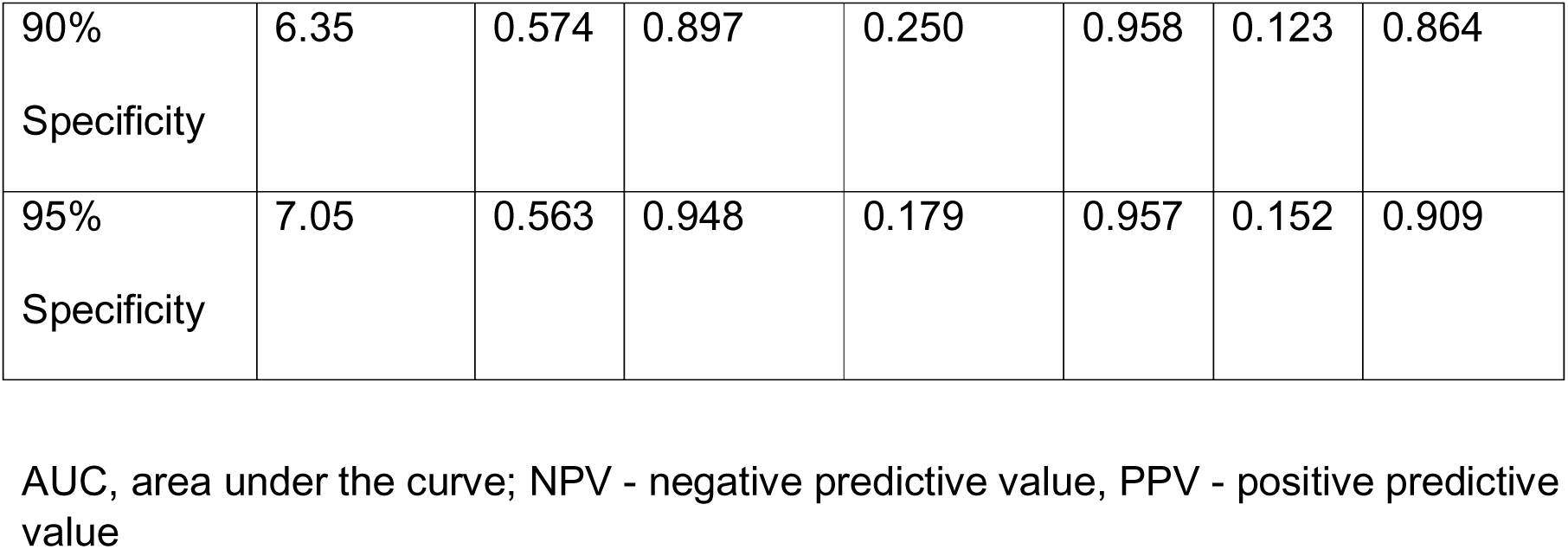
Diagnostic performance of selected ECG LVH criteria at different cut-off values.

**Figure S1.**
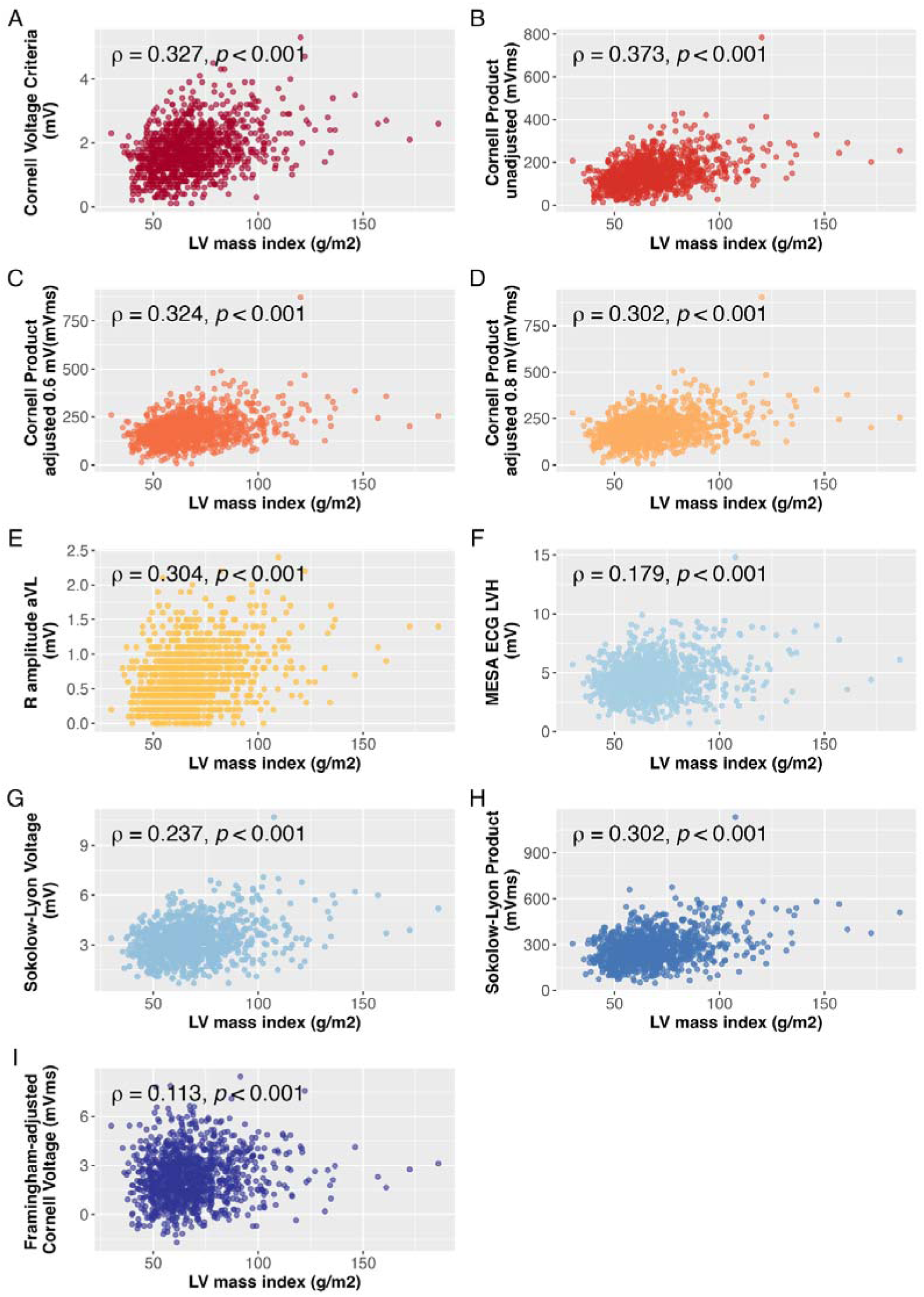
Scatter plots showing correlation between continuous ECG parameters and LV mass index from TTE parameters. Panel A: Cornell voltage criteria; Panel B: unadjusted Cornell voltage product; Panel C: Cornell voltage product, adjusted for women 0.6 mV; Panel D: Cornell voltage product, adjusted for women 0.8 mV; Panel E: R amplitude in lead aVL; Panel F: General MESA ECG-LVH; Panel G: Sokolow-Lyon voltage; Panel H: Sokolow-Lyon product; Panel I: Framingham-adjusted Cornell voltage

**Figure S2:**
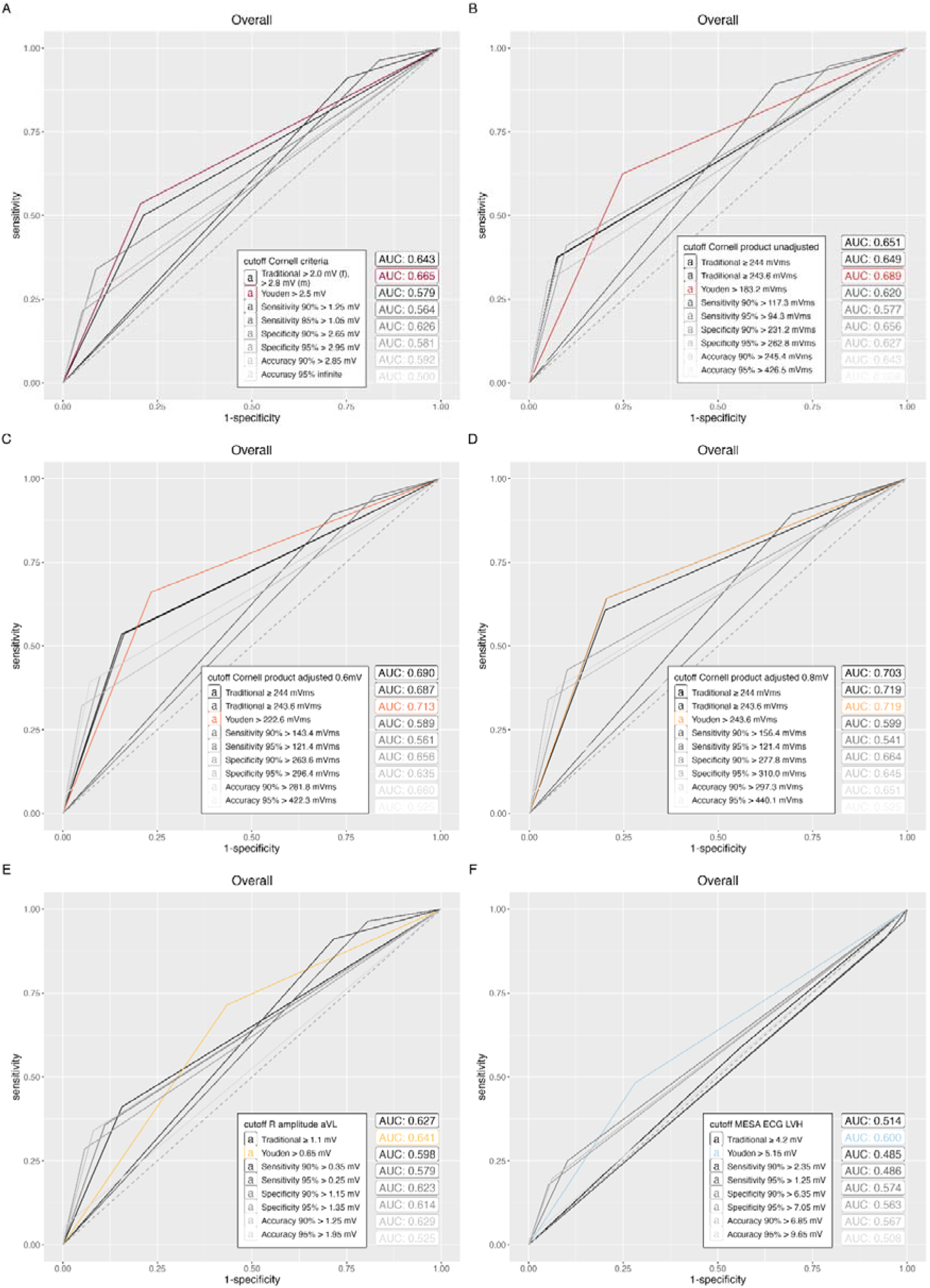
Comparison of AUCs for traditional vs. Youden-index-derived cut-offs, and AUCs at sensitivity, specificity, or accuracy levels exceeding 90% and 95%.

